# Social and racial inequalities in COVID-19 risk of hospitalisation and death across São Paulo state, Brazil

**DOI:** 10.1101/2020.12.09.20246207

**Authors:** Sabrina L Li, Rafael H M Pereira, Carlos A Prete, Alexander E Zarebski, Lucas Emanuel, Pedro JH Alves, Pedro S Peixoto, Carlos KV Braga, Andreza A de Souza Santos, William M de Souza, Rogerio J Barbosa, Lewis F Buss, Alfredo Mendrone, Cesar de Almeida-Neto, Suzete C Ferreira, Nanci A Salles, Izabel Marcilio, Chieh-Hsi Wu, Nelson Gouveia, Vitor H Nascimento, Ester C Sabino, Nuno R Faria, Jane P Messina

**Affiliations:** School of Geography and the Environment, University of Oxford, Oxford, United Kingdom; Institute for Applied Economic Research, Brasília, Brazil; Department of Electronic Systems Engineering, University of São Paulo, São Paulo, Brazil; Department of Zoology, University of Oxford, Oxford, United Kingdom; Department of Applied Mathematics, Institute of Mathematics and Statistics, University of São Paulo, São Paulo, Brazil; Oxford School of Global and Area Studies, Latin American Centre, University of Oxford, Oxford, United Kingdom; Virology Research Center, University of São Paulo, Ribeirão Preto, Brazil; Center for Metropolitan Studies, Faculty of Philosophy, Languages and Literature, and Human Sciences, University of São Paulo, São Paulo, Brazil; Departamento de Molestias Infecciosas e Parasitarias & Instituto de Medicina Tropical da Faculdade de Medicina da Universidade de São Paulo, São Paulo, Brazil; Fundação Pró-Sangue Hemocentro de São Paulo; Disciplina de Ciências Médicas, Faculdade de Medicina da Universidade de São Paulo, São Paulo, Brazil; Laboratory of Medical Investigation in Pathogenesis and Directed Therapy in Onco – Immuno – Hematology (LIM-31) HCFMUSP, University of São Paulo Medical School, São Paulo, Brazil; Epidemiologic Surveillance Center, Hospital das Clinicas – University of São Paulo Medical School, São Paulo, Brazil; Mathematical Sciences, University of Southampton, Southampton, United Kingdom; Department of Preventive Medicine, University of São Paulo Medical School; MRC Centre for Global Infectious Disease Analysis, J-IDEA, Imperial College London, London, UK; Oxford School of Global and Area Studies, University of Oxford, Oxford, United Kingdom

## Abstract

**Background:** Little evidence exists on the differential health effects of COVID-19 on disadvantaged population groups. Here we characterise the differential risk of hospitalisation and death in São Paulo state, Brazil and show how vulnerability to COVID-19 is shaped by socioeconomic inequalities.

**Methods:** We conducted a cross-sectional study using hospitalised severe acute respiratory infections (SARI) notified from March to August 2020, in the *Sistema de Monitoramento Inteligente de São Paulo* (SIMI-SP) database. We examined the risk of hospitalisation and death by race and socioeconomic status using multiple datasets for individual-level and spatio-temporal analyses. We explained these inequalities according to differences in daily mobility from mobile phone data, teleworking behaviour, and comorbidities.

**Findings:** Throughout the study period, patients living in the 40% poorest areas were more likely to die when compared to patients living in the 5% wealthiest areas (OR: 1·60, 95% CI: 1·48 – 1·74) and were more likely to be hospitalised between April and July, 2020 (OR: 1·08, 95% CI: 1·04 – 1·12). Black and *Pardo* individuals were more likely to be hospitalised when compared to White individuals (OR: 1·37, 95% CI: 1·32 – 1·41; OR: 1·23, 95% CI: 1·21 – 1·25, respectively), and were more likely to die (OR: 1·14, 95% CI: 1·07 – 1·21; 1·09, 95% CI: 1·05 – 1·13, respectively).

**Interpretation:** Low-income and Black and *Pardo* communities are more likely to die with COVID-19. This is associated with differential access to healthcare, adherence to social distancing, and the higher prevalence of comorbidities.

**Funding:** This project was supported by a Medical Research Council-São Paulo Research Foundation (FAPESP) CADDE partnership award (MR/S0195/1 and FAPESP 18/14389-0) (http://caddecentre.org/). This work received funding from the U.K. Medical Research Council under a concordat with the U.K. Department for International Development.

## Introduction

The COVID-19 pandemic has amplified the effects of social inequalities on exposure and death in low socioeconomic groups ^1^, particularly in Brazil, where it has caused significant mortality ^2^. Few studies have addressed the uneven impact of COVID-19 by socioeconomic status and race ^3–5^, especially in low and middle-income countries, in part because national surveillance systems seldom collect or report this information ^6^. In Brazil, higher risk of COVID-19 death has been found for Black and *Pardo* (mixed ethnicity) Brazilians ^7^. Nonetheless, there is still little information on how the differential health outcomes of COVID-19 are shaped by broader social inequalities that determine the capacity to adhere to social distancing and non-pharmaceutical interventions (NPIs).

It is paramount to understand the potential social drivers of COVID-19 morbidity and mortality, particularly in countries with high inequality such as Brazil ^8^. The first COVID-19 cases in Brazil were detected in São Paulo ^9^, the most populous state and home to diverse racial groups. In the Brazilian context of politically polarised public health responses ^10^, São Paulo has been severely affected by COVID-19 ^11^ and access to testing has been limited for low-income populations ^12^. By combining multiple high-resolution data sources, we conducted a multi-scale analysis to investigate the risk of hospitalisation and death from severe acute respiratory infections (SARI), predominantly caused by COVID-19 ^12^, by race and socioeconomic status in the state of São Paulo. We examined potential drivers of these inequalities by evaluating local levels of population adherence to social distancing, access to teleworking, and prevalence of comorbidities

## Methods

### Data Sources

#### Severe acute respiratory infections (SARI) and patient information

Patient-level information on demographic characteristics, home address, hospitalisation, and health outcomes were collected from the São Paulo State Health Secretariat SARI hospitalisations database (SIMI-SP) ^13^. SARI can be caused by SARS-CoV-2 and is defined by the Brazilian Ministry of Health as flu-like syndrome plus one of the following: dyspnoea, persistent chest pain, or hypoxia. All SARI cases and deaths are notified in the SIMI-SP database, regardless of hospitalisation.

We included all SARI related hospitalisations and deaths notified in São Paulo state between March 15 and August 29, 2020. Given that recent data is incomplete due to reporting delays ^14^ and to avoid biases, we limited our analysis to patients with symptoms onset between these dates (epidemiological weeks 10 – 35) (figure 1B). We also included SARI cases with unknown aetiology, as those are likely related to COVID-19 but not lab-confirmed due to low rates of COVID-19 testing in Brazil ^15^ and socioeconomic bias in testing ^12^.

**Figure 1.**
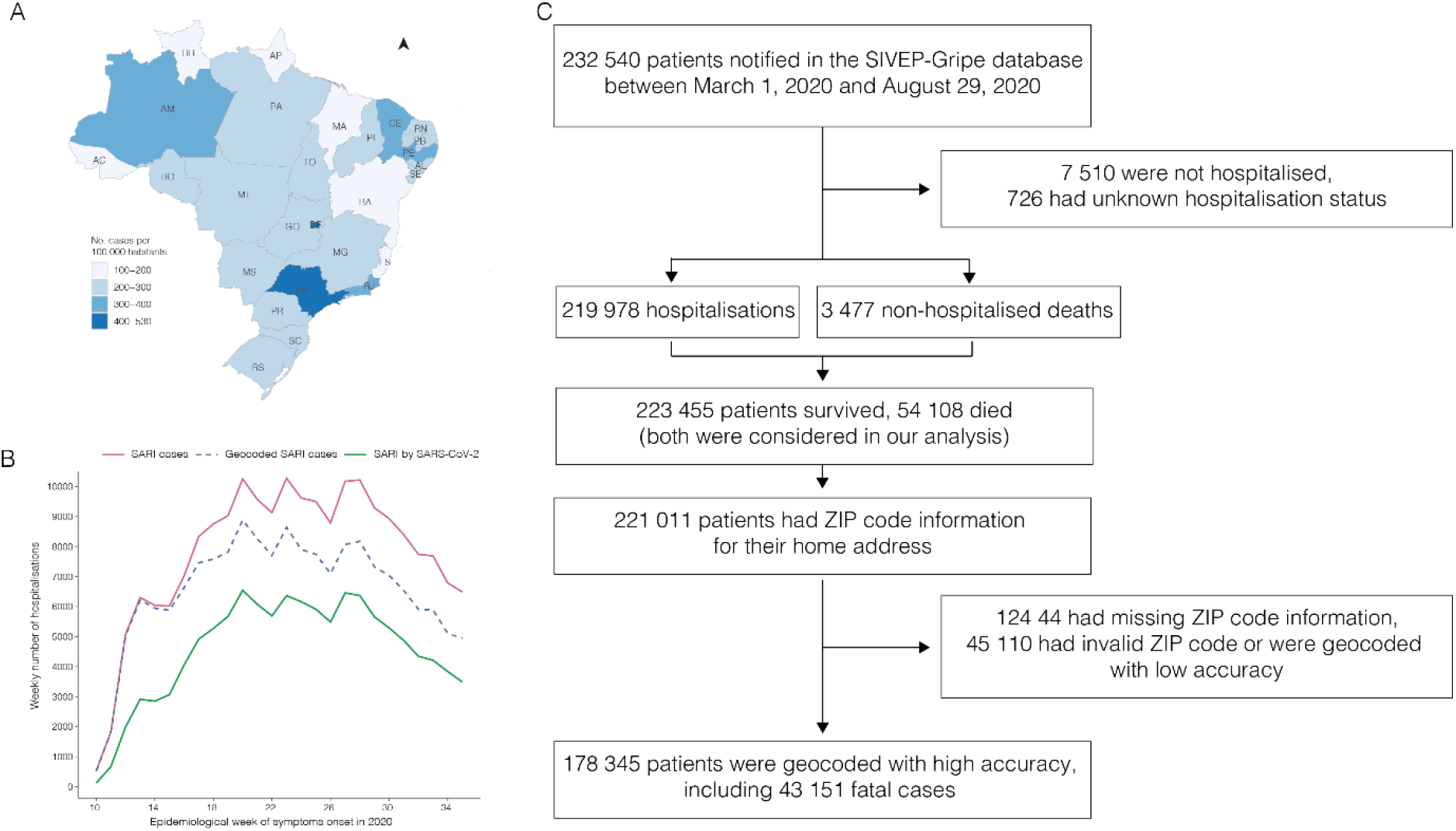
Number of SARI hospitalisations per 100 000 habitants by state in Brazil between March 1 and August 29, 2020 (A). Number of SARI hospitalisations for the state of São Paulo and the greater metropolitan area of São Paulo (RMSP) by week of symptom onset (B). Flowchart of SIMI-SP data processing (Source: https://covid.saude.gov.br) (C).

Zip code information was only available for cases reported in São Paulo state. Data was geocoded using the patient’s self-reported home address or postal code with Galileo (www.img.com.br) and Google API. Information on the health facility where each case was notified was linked to the National Registry of Health Facilities (CNES), which includes information on the mode of health care provision (public and private).

The race of patients was partially self-declared and partially identified by a health professional. Race was categorised as either “White”, “Black”, “Asian” (East or Southeast Asian), “*Pardo”* (mixed ethnic ancestries with diverse skin colours)^16^ or Indigenous. Race information was missing for 53 480 (23·9%) of retrieved SARI cases, and were imputed using the racial distribution of the census tract of residence (see Supplementary Appendix A for details). About 0·1% of the population in São Paulo self-identified as indigenous ^17^ and since only 166 indigenous patients (0·07%) were recorded, they were not considered in our analysis.

#### Socioeconomic data

We obtained data on municipality-level socioeconomic factors from the latest population census (2010) compiled by the Brazilian Institute of Geography and Statistics ([IBGE](https://www.ibge.gov.br/estatisticas/sociais/populacao/9662-censo-demografico-2010.html?=&t=o-que-e)). We selected household income per capita, population density, and income inequality (Gini Index). We also determined the proportion of residents with a primary education or lower, employment-to-population ratio, and the proportion of the working population without a formal labour market contract or social security. The road network distance from the centroid of each census tract to the nearest healthcare facility was computed considering all facilities that hospitalised SARI patients via the public healthcare system (SUS). Information on employment status and comorbidities during the epidemic was retrieved from the National Household Sample Survey (PNAD COVID-19), a national telephone survey conducted by IBGE with over 1 888 560 interviews between May and September 2020. Details are described in Supplementary Appendix A.

#### Daily mobility and non-pharmaceutical interventions

To assess local-level adherence to social distancing, we used daily mobile phone data provided by In-Loco (https://www.inloco.com.br/covid-19)^18^ for the *Região Metropolitana de São Paulo* (greater metropolitan area of São Paulo: RMSP). This data was aggregated using a hexagonal grid based on the global H3 index at resolution 8. Each cell has an edge of approximately 460 meters and an area of 0·74 km^2^ (https://h3geo.org/docs/core-library/restable). For each H3 cell, the social isolation index was measured as the number of people who did not leave their cell of residence during the day, divided by the number of residents in that cell. Each mobile phone was assigned to an H3 cell based on the owner’s location of residence during the evening and their previous travel history. The racial composition and income level of each cell were determined using dasymetric interpolation (Supplementary Appendix A).

We also used municipality and state-level data on NPIs from a continuous survey conducted between May 13 and July 31, 2020 ^19^. The survey had 13 questions related to the implementation and easing of social distancing measures, and responses were obtained from 612 mayors in São Paulo (94·8% of the total).

## Data analysis

### Probability of hospitalisation and death

We conducted an individual-level analysis to estimate the probability of reporting a SARI hospitalisation given a patient’s race and average income level in their census tract of residence. Census tracts were grouped by quantiles of income per capita into six categories as presented in the results. Similarly, we determined the probability of death from SARI given a patient’s race, income, and administrative type of the health facility where the patient was hospitalised (public or private). Both probabilities were standardised by age and sex to account for demographic differences between groups. They were calculated for every month between March and August 2020. Probabilities for each age-sex group were estimated empirically using relative frequencies. Odds ratios using White patients and the highest income level as reference groups were computed. Confidence intervals were calculated using bootstrapping. Details are in Supplementary Appendix A.

### Seroprevalence by socioeconomic status

To assess the broader risk of SARS-CoV-2 infection beyond hospitalisation, we analysed seroprevalence data collected as part of the national Covid-IgG study ^20^ from blood donors aged 16 – 69 living in São Paulo city ^20^ (Supplementary Appendix A). We calculated the proportion of individuals by education and race category with detectable anti-SARS-CoV-2 antibodies between February and October 2020.

### Socioeconomic drivers and hospitalisation risk

An ecological spatio-temporal regression analysis was conducted at the municipality level for São Paulo state (n = 645 municipalities) to assess the monthly risk of hospitalisation and its association with socioeconomic factors between March 1 and August 29, 2020. To further understand the association between socioeconomic conditions and COVID-19 risk, we conducted the same analysis at the census tract level (n= 30 815) for the RMSP, where the majority of cases were concentrated. The relative risk of hospitalisation was estimated using a hierarchical Bayesian model composed of a generalised log-linear model with spatially structured and unstructured random effects to account for spatial autocorrelation and time-varying random effect. The spatial structure is characterised by population movement between municipalities from March 1 to August 15, 2020 defined by In-Loco mobile geo-location data summarised elsewhere ^18^. A detailed description of the model and interpretation, covariates, and diagnostics can be found in Supplementary Appendix A.

### Social adherence response to NPIs

We used an event study design ^21^ to examine how different socioeconomic groups changed their daily mobility levels in response to the introduction and relaxation of NPIs in the RMSP. We compared changes in mobility patterns of the population living in H3 cells with predominantly White versus predominantly Black or *Pardo* residents, as well as of the population living in areas of the wealthiest and poorest income quintiles. The daily social distancing index from hexagons were regressed on a set of relative time dummies that indicated the number of days before and after the first NPI introduction in São Paulo state. Hexagon-fixed effects controlled for unobserved time-invariant determinants of social distancing, while day-fixed effects controlled for temporal shocks common to all hexagons. We further included an additional time-varying control variable, representing the number of days relative to the first confirmed SARI case in each hexagon, and a dummy variable indicating the period of NPI relaxation in each municipality. Sensitivity analyses were performed and discussed in Supplementary Appendix A.

We employed a multinomial logistic regression to estimate the probability that employed individuals would be working face-to-face, teleworking, or taking paid or unpaid leave. Differences in the work status of individuals by race, education, and occupation were calculated while controlling for age and sex. 35 groups of employment occupations listed in the PNAD COVID-19 survey were aggregated to ten SCO-08 1-digit occupational groups defined by the International Labour Organisation. We further disaggregated health professionals and health technicians (Supplementary Appendix A – Table A2).

### Comorbidities

PNAD COVID-19 data was used to conduct a binomial logistic regression to estimate the odds ratio of having at least one comorbidity, by race and education attainment (pre-primary, primary, secondary, and tertiary), while controlling for age and sex for São Paulo state. The comorbidities considered were chronic obstructive pulmonary disease (COPD), diabetes, hypertension, or cardiovascular disease such as myocardial infarction, angina or heart failure. Confidence intervals for the odds ratios were calculated taking into account PNAD’s complex sample design.

## Results

Between March 1 and August 29, 2020, São Paulo state had the highest number of SARI hospitalisations per 100 000 habitants compared to all states in Brazil (figure 1A). A time series illustrating the number of SARI hospitalisations is presented in figure 1B. During this time, 232 540 patients were notified in the SIMI-SP database (figure 1C), from which 230 794 (99·2%) had a confirmed COVID-19 diagnosis or were diagnosed with SARI of unknown or missing aetiology. From these, 223 455 were hospitalised (98·4%) or died without hospitalisation (1·6%). From the non-hospitalised cases, we only selected deaths; 54 108 patients died, of which 52·5% were White, 20·2% were *Pardo*, 6·13% were Black, 1·96% were Asian, 0·052% were Indigenous and 19·6% did not have race information. We geocoded 178 345 (79·8%) of all SARI cases considered in our analyses with high accuracy at either street address, route, or neighbourhood level without compromising personal privacy.

During the first month of the COVID-19 epidemic (March) in Brazil, hospitalised patients were more likely to be White or Asian and come from census tracts with higher income per capita (figure 2A and 2B). During this period, people living in low-income areas, compared to high-income areas, were less likely (OR: 0·44, 95% CI: 0·42 – 0·46) to be hospitalised with SARI. This coincides with the early establishment of COVID-19 and its first cases in higher-income travellers returning from overseas ^9,12^.

**Figure 2.**
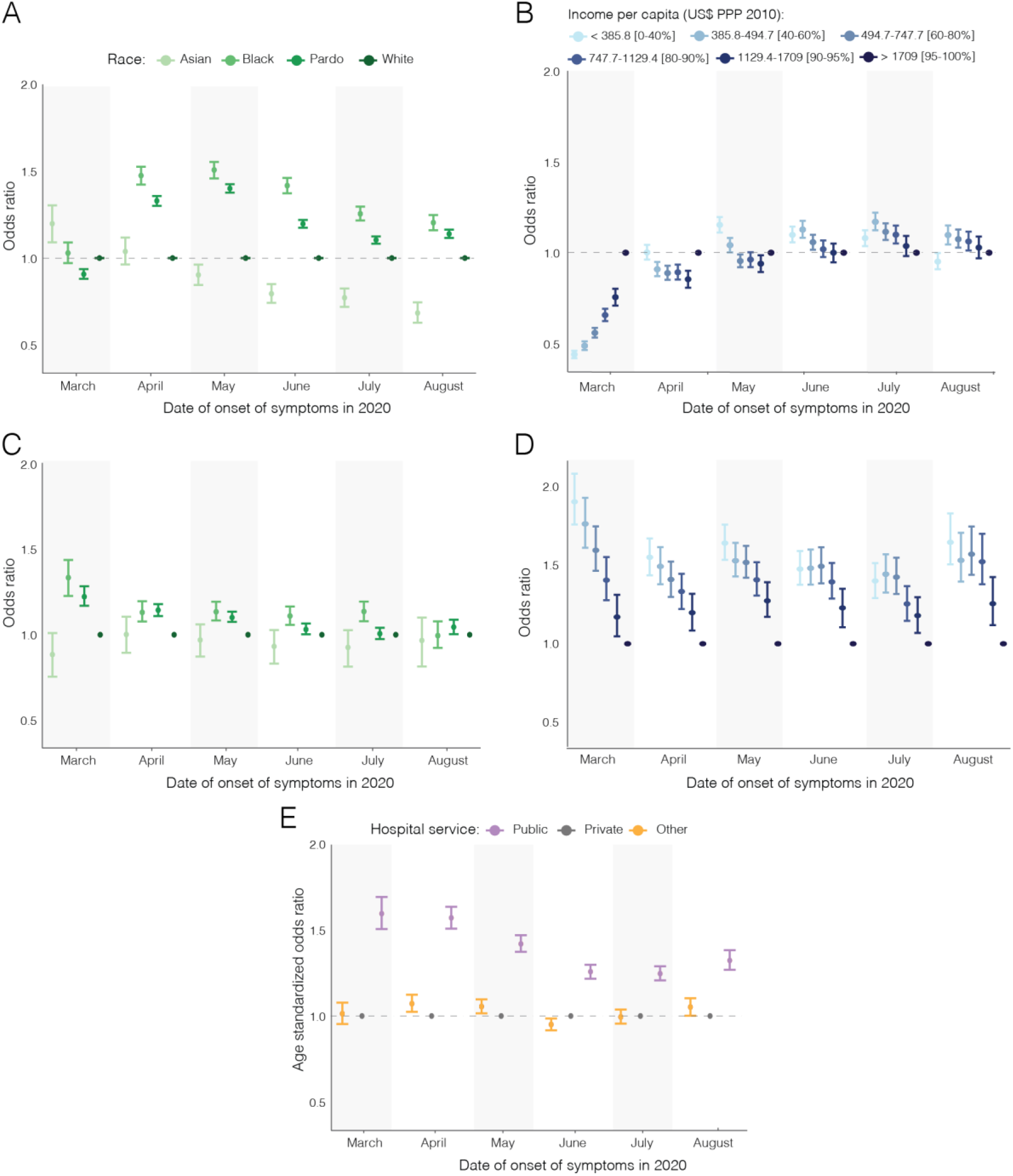
Age-standardised odds ratio (OR) for SARI hospitalisation by race (A) and by income (B); Age-standardised OR of death among SARI cases by race (C) and income (D); and Age-standardised OR of death among SARI cases by hospital type for São Paulo state (E). *PPP: Purchasing power parity*.

As the epidemic progressed from April onwards, patients were also more likely to be from low-income census tracts (April to July, OR: 1·08, 95% CI: 1·04 – 1·12), except for August, when patients were less likely to be from low-income census tracts (OR: 0·95, 95% CI: 0·91 – 1·00). Similarly, Black Brazilians and *Pardos* became more likely to be hospitalised with SARI than Whites (OR: 1·37, 95% CI: 1·32 – 1·41; OR: 1·23, 95% CI: 1·21 – 1·25, respectively), while Asians became the least likely to be hospitalised (OR: 0·90, 95% CI: 0·83 – 0·96). These results were further confirmed by our seroprevalence findings, which showed that anti-SARS-CoV-2 antibodies were highest in Black blood donors and those with low educational attainment across all age groups (Supplementary Appendix B – figure S1).

Once hospitalised, Black and *Pardo* patients were more likely to die from SARI than White patients (OR: 1·14, 95% CI: 1·07 – 1·21; 1·09, 95% CI: 1·05 – 1·13, respectively) (figure 2C). This difference was more pronounced in March and was reduced over time. We found that patients living in the poorest census tracts were more likely to die from SARI compared to patients from the wealthiest tracts (OR: 1·60, 95% CI: 1·48 – 1·74) (figure 2D). Likewise, patients treated in public hospitals were more likely to die than patients treated in private hospitals throughout the epidemic (OR: 1·40, 95% CI: 1·34-1·46) (figure 2E). Racial differences in the probability of death decreased when considering only patients hospitalised at public health facilities but persisted among patients in private facilities (Supplementary Appendix B – figure S2).

To understand the geographical variation in SARI hospitalisation, we predicted and mapped the relative risk of SARI hospitalisation for São Paulo state by month via a model with a spatial structure defined by human movement fluxes derived from anonymised mobile phone data (figure 3A) and covariates related to socioeconomic status (figure 3B). This is shown in figure 3C. We found a lower risk of hospitalisation in municipalities with high income per capita (fixed effect = −0·93, 95% CI: −1·17 – −0·69) and high proportion of adult residents with a primary education or lower (−0·80, 95% CI: −1·0 – −0·60). Municipalities with fewer nearby public health facilities were also found to have lower risk of hospitalisation (−0·24, 95% CI: −0·45 – −0·03). We also found a higher risk of SARI hospitalisation in municipalities with higher population density (0·24, 95% credible interval: 0·04 – 0·44).

**Figure 3:**
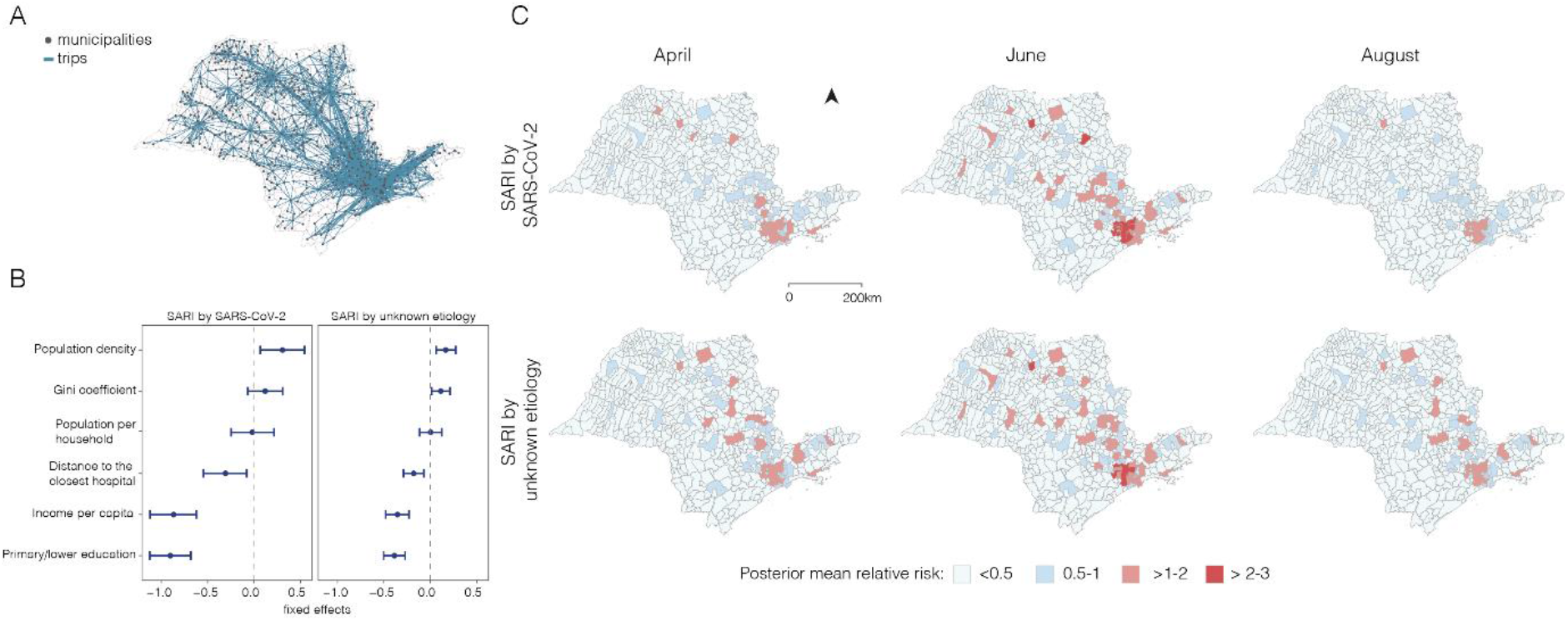
Relative risk of SARI hospitalisation in the state of São Paulo at the municipality level (A). Human movement between municipalities based on In-Loco mobile phone data retrieved from March to August 2020 (B). Fixed effects and 95% credible intervals for socioeconomic covariates (C).

We found that over time, the risk of SARI hospitalisation increased particularly in municipalities near and within the RMSP, where 70% of the SARI cases reported for the state are concentrated (figure 4A). By mapping risk at the census tract level for the RMSP, we detected increasing risk starting from São Paulo city (central region). By June, almost all of the census tracts in and near the city centre were found to have high relative risk, but this risk decreased by August.

**Figure 4.**
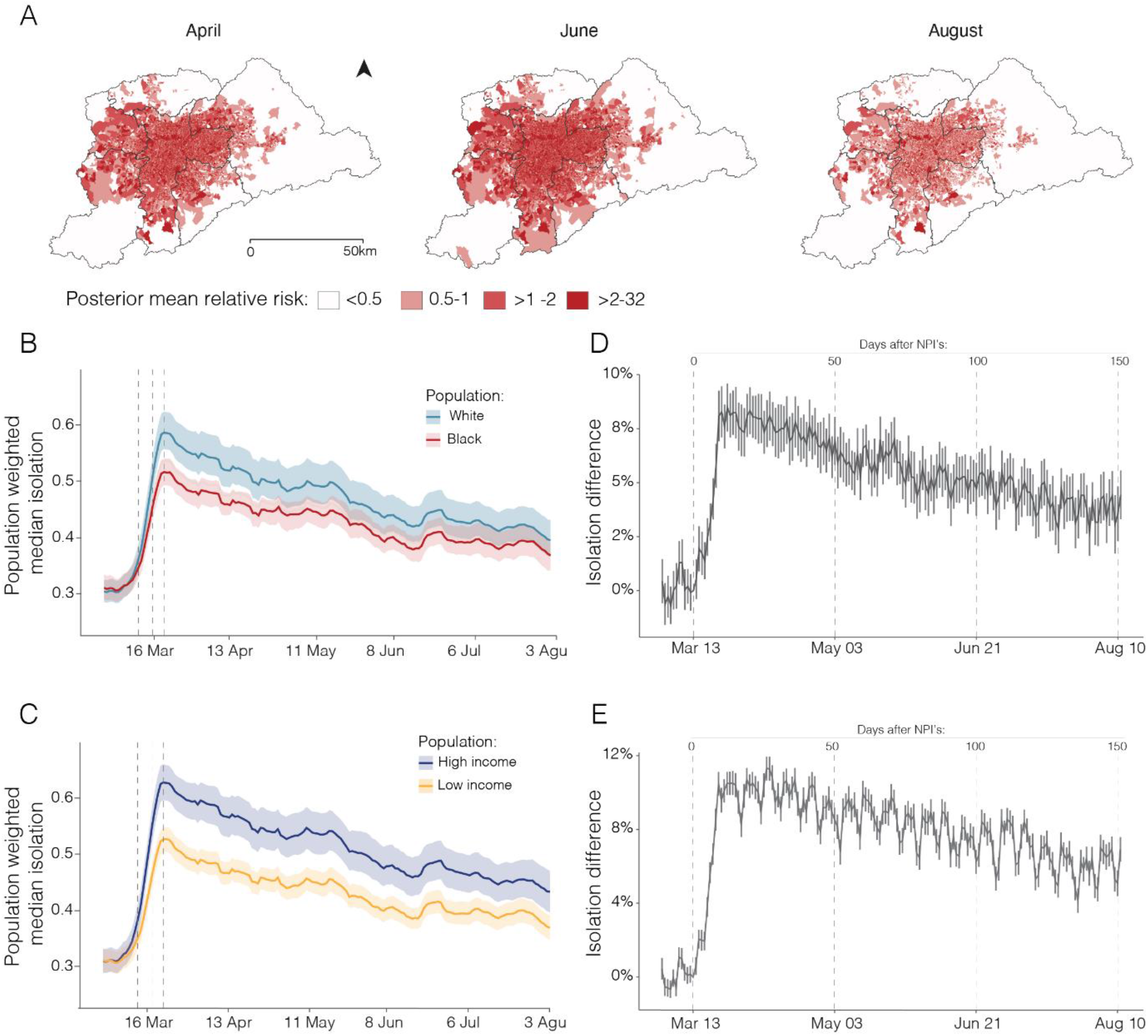
Relative risk of SARI hospitalisation for the RMSP (A). Seven-day moving average of daily median levels in social distancing by race (B) and income (C). Difference in daily social isolation by race (D) and income (E) after the introduction of NPIs. In panels (B) and (C), solid lines show population-weighted median isolation levels and shaded areas show population-weighted interquartile range (25% - 75%). Dashed vertical lines indicate the dates of NPIs that enabled school closure (March 13 was the state NPI) and non-essential activities (March 18 and 22, municipal and state NPIs, respectively).

Differential risk to SARI in the RMSP was also associated with daily mobility levels by disadvantaged groups. Before the implementation of NPIs on March 13, mobility levels were similar across all socioeconomic groups (figures 4B and 4C). However, 14 days after the introduction of NPIs, isolation levels were 8·2% (95% CI: 7·2% – 9·2%) higher in predominantly White areas compared to predominantly Black areas. Similarly, 27 days after the introduction of state-level NPIs, isolation levels were 11·2% (95% CI: 10·6% – 11·9%) higher in the wealthiest than in the poorest areas. Overall, we detected a decreasing trend in isolation levels over time, and the magnitude of the differences in social isolation levels between areas with predominantly White and Black populations decreased to only 4·4% (95% CI: 3·3% – 5·5%) 151 days after the introduction of the NPIs (figure 4D).

Finally, we investigated the differential risk to SARI based on workers’ position in the labour market using data from the PNAD COVID-19 survey. After the introduction of NPIs, workers employed in low-skilled jobs or essential services were more likely to keep working face-to-face than workers in professional or managerial positions (Supplementary Appendix B - figure S3). Workers with pre-primary education were more likely to work in occupations that require in-person contact than workers with tertiary education (Probability: 0·89, 95% CI: 0·87 – 0·90 compared to PR: 0·58, 95% CI: 0·57 – 0·60) and less likely to work in occupations that allow teleworking (PR: 0·005, 95% CI: 0·004 – 0·007 versus PR: 0·36 95% CI: 0·35 – 0·37, respectively) (figure 5A). When controlling for education and formal or informal employment, we found no substantial difference between racial groups in the probability of working face-to-face or teleworking (figure 5B).

**Figure 5:**
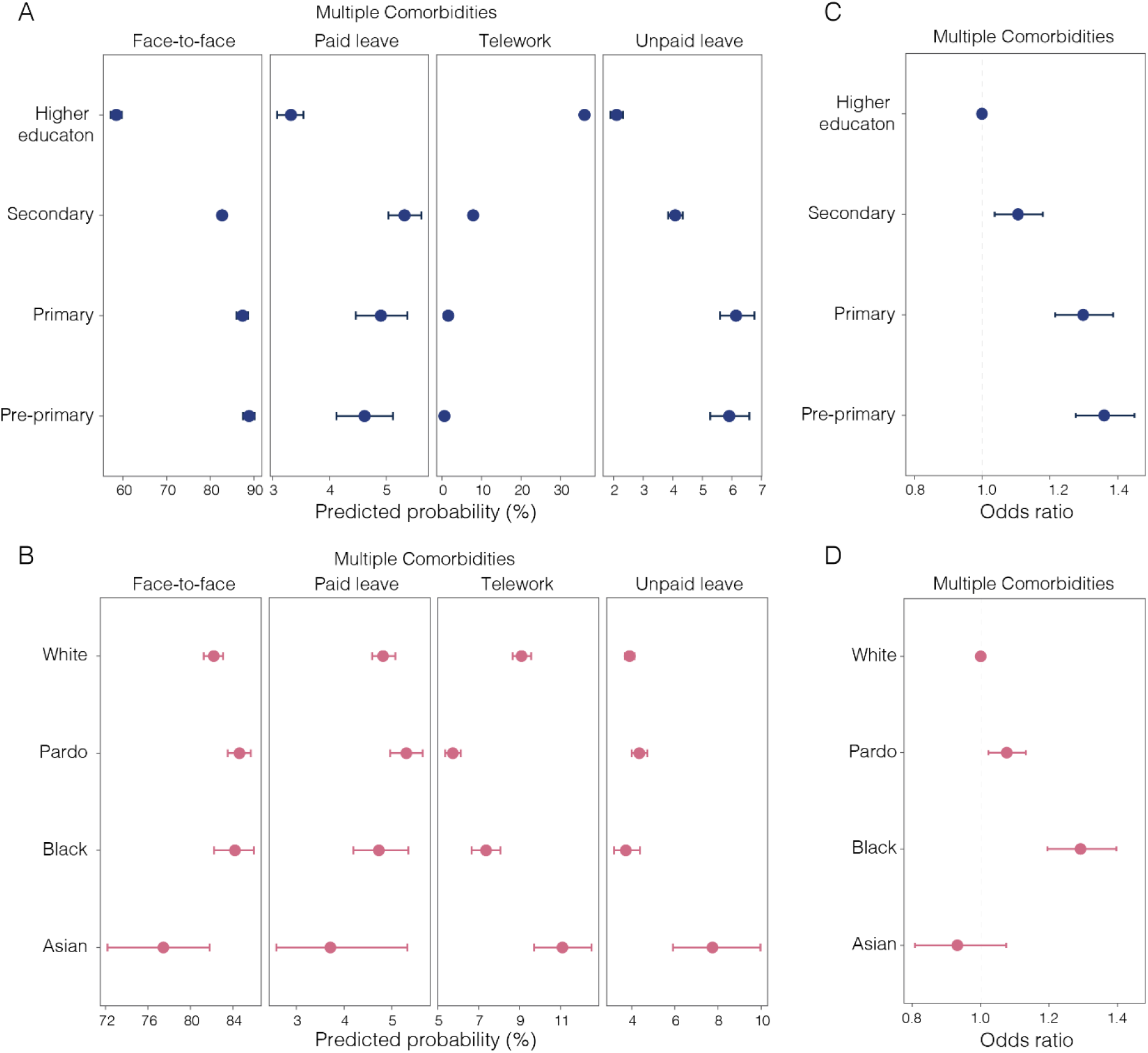
Probability of different working conditions by education attainment (A) and race (B). Odds ratio (OR = 1) of having multiple comorbidities: chronic obstructive pulmonary disease (COPD), diabetes, hypertension or cardiovascular disease such as infarction, angina, and heart failure by education attainment (C) and race (D). Horizontal lines show 95% confidence intervals. Source: PNAD COVID-19 /IBGE ^17^, July to September, 2020.

We found that population groups at risk of death from SARI were also more likely to have comorbidities. Compared to the population with tertiary education in São Paulo state, individuals with primary education or lower are more likely to have one or more comorbidities (OR: 1·36, 95% CI: 1·27 – 1·45) (figure 5C). Similarly, Black individuals were also more likely to have one or more comorbidities than White individuals (OR: 1·29, 95% CI: 1·19 – 1·39) (figure 5D). Odds ratio estimates for each health condition are summarised in Supplementary Appendix B – figure S4.

## Discussion

Our study shows that disadvantaged hospitalised groups are disproportionately more likely to die from SARI. We find that the differential health outcomes can be explained by structural inequities linked to the incidence of comorbidities and to socioeconomic status, which limit the ability of these groups to socially isolate and reduce their access to optimal health services.

Social and racial inequalities shape the risk of SARI hospitalisation and death. After the initial phase of international imports in Brazil ^12^, Black or *Pardo* Brazilians residing in low-income areas were more likely to be hospitalised and die with SARI compared to White individuals, which aligns with recent findings ^7^. Our assessment of anti-SARS-CoV-2 antibodies in blood donors categorised by demographic background further confirms that Black Brazilians and those with lower socioeconomic status are disproportionately exposed to COVID-19. While patients hospitalised in public health facilities were more likely to die than those in private health facilities, racial inequalities in death probability are attenuated when only considering patients within hospitals of the same type, either public or private. Potential factors influencing this inequality include higher comorbidities among poor Black patients and the lower access to private care among low-income individuals that are disproportionately Black.

We found higher hospitalisation risk for those living in municipalities with low income per capita and high population density compared to the rest of São Paulo state. These populations mainly reside in the RMSP, which contains nearly half of the population in São Paulo state and where bias in testing is evident in regions of lower socioeconomic status ^12^ (Supplementary Appendix B – figure S5). The risk of SARI hospitalisation is particularly elevated in São Paulo city, where seroprevalence estimates from blood donors show that anti-SARS-CoV-2 antibodies were highest in older Black Brazilians and those with lower educational attainment.

We show that inequalities in the risk of SARI hospitalisation were partially explained by differential mobility responses to social distancing guidelines, similar to the US ^22^. In wealthier and predominantly White neighbourhoods, people were able to isolate faster and sustain isolation for long periods of time. Among the working population, low-income and Black workers were less likely to receive a furlough from work or telework. Due to systemic inequalities in education and the labour market, these groups are disproportionately employed in precarious job positions with no social security and dependent on day-to-day income ^23^, limiting their ability to reduce mobility. Other important factors include the disadvantage of having multiple comorbidities, which are more prevalent among Blacks and *Pardos* and those with lower education.

Our study has limitations that may have underestimated the level of inequality. Firstly, geocoding cases may have discarded patients from poor census tracts where accuracy is limited ^24^. Secondly, using data aggregated for various administrative levels has inherent limitations due to ecological fallacy and the modifiable areal unit problem ^25^. Finally, the 2010 Brazilian population census and PNAD COVID-19 survey may have limited the capture of socioeconomic changes in the last decade and inclusion of extremely wealthy individuals ^26^. Additionally, disadvantaged groups can be underrepresented in health administrative records because of their lower access to healthcare. Given that São Paulo is the wealthiest state and has the most robust healthcare system in Brazil ^27^, it is possible that the impact of inequalities is more severe in other states.

Our findings on the difference in risk of SARI death reveal stark inequalities in access to healthcare. Only 25% of Brazilians have access to private healthcare via health insurance, reflecting how inequality in access to quality healthcare is largely driven by income ^28^. This leaves 75% of the population solely reliant on a chronically underfunded public healthcare system, which highlights that disadvantaged populations are more likely to be infected and deprived of care. Strengthening healthcare access and its capacity will be critical for reducing health inequities during this and forthcoming public health emergencies ^29^.

Our findings on socioeconomic risk factors could help guide vaccine allocation in diverse settings to achieve equitable access. Ensuring that disadvantaged groups, especially those that have in-person occupations and live in crowded and deprived areas receive vaccination will help prevent and slow down community transmission.

While race is not a risk factor in itself, it is critical to consider systemic inequalities that lead Black and *Pardo* communities to be overrepresented among low socioeconomic groups, to have higher rates of severe COVID-19 infection, and comorbidities that exacerbate their risk of death. Therefore, including disadvantaged populations among priority groups for vaccination could help reduce health inequities instead of exacerbating them ^30^.

Our study highlights the need for additional research to comprehend the effects of social and health inequalities during pandemics. Firstly, an assessment of the inequality in access to quality care within public and private health facilities and its risk factors is needed to better understand mortality in different hospital settings. Secondly, our study shows that population response to NPIs can vary significantly based on social circumstances, suggesting that future studies should also consider socioeconomic aspects when evaluating the effectiveness of NPIs. Thirdly, more data is needed on whether social safety net programs that are guaranteeing income for disadvantaged groups during the pandemic (e.g., Brazil’s emergency cash payment), may have enabled people to reduce their mobility. Nevertheless, our study has shown the impact of social inequities on COVID-19 hospitalisation and death, thus informing future research and policies related to the health impacts of COVID-19 in Latin America.

## Data Availability

The data sets used in the research are available from the corresponding authors upon request and ethical approval.

## Contributors

SLL and RHMP conceived the research questions and designed the study. RHMP, CAPJ, LE, PJHA, PSP, CKB, AASS, WMS, RJB, LFB, AM, CAN, SCF, and NAS collected the epidemiological, socioeconomical, mobility, NPIs, seroprevalence, and comorbidities data. SLL, RHMP, CAPJ, AEZ, LE, PJHA, CKB, RJB, and VN conducted exploratory and statistical analysis and interpreted the results. SLL, RHMP, and CAPJ wrote the manuscript. All authors read and revised the final manuscript.

## Acknowledgement

SLL is supported by the Oxford Martin School and the Canadian Social Sciences and Humanities Doctoral Fellowship. CAPJ is supported by Coordenação de Aperfeiçoamento de Pessoal de Nível Superior - Brasil (CAPES) and Fundação Faculdade de Medicina (FFM). AEZ is supported by The Oxford Martin Programme on Pandemic Genomics. WMS is supported by the São Paulo Research Foundation, Brazil (2017/13981-0 and 2019/24251-9). NRF is supported by a Wellcome Trust and Royal Society Sir Henry Dale Fellowship (204311/Z/16/Z).

## References

1 Ahmed F, Ahmed N, Pissarides C, Stiglitz J. Why inequality could spread COVID-19. Lancet Public Health 2020; 5: e240.

2 Walker PGT, Whittaker C, Watson OJ, et al. The impact of COVID-19 and strategies for mitigation and suppression in low- and middle-income countries. Science 2020; 369: 413–22.

3 Drefahl S, Wallace M, Mussino E, et al. A population-based cohort study of socio- demographic risk factors for COVID-19 deaths in Sweden. Nat Commun 2020; 11: 5097.

4 UK Office for National Statistics. Coronavirus (COVID-19) related deaths by ethnic group, England and Wales - Office for National Statistics. 2020https://www.ons.gov.uk/peoplepopulationandcommunity/birthsdeathsandmarriages/deaths/articles/coronavirusrelateddeathsbyethnicgroupenglandandwales/2march2020to10april2020 (accessed Nov 20, 2020).

5 Cummings MJ, Baldwin MR, Abrams D, et al. Epidemiology, clinical course, and outcomes of critically ill adults with COVID-19 in New York City: a prospective cohort study. The Lancet 2020; 395: 1763–70.

6 Pan D, Sze S, Minhas JS, et al. The impact of ethnicity on clinical outcomes in COVID-19: A systematic review. EClinicalMedicine 2020; 23: 100404.

7 Baqui P, Bica I, Marra V, Ercole A, Schaar M van der. Ethnic and regional variations in hospital mortality from COVID-19 in Brazil: a cross-sectional observational study. Lancet Glob Health 2020; 8: e1018–26.

8 Facundo A, Chancel, Lucas, Piketty, Thomas, Saez, Emmanuel, Zucman, Gabriel. World Inequality Report 2018. 2017.

9 Jesus JG de, Sacchi C, Candido D da S, et al. Importation and early local transmission of COVID-19 in Brazil, 2020. Rev Inst Med Trop São Paulo 2020; 62: e30.

10 Henriques CMP, Vasconcelos W, Henriques CMP, Vasconcelos W. Crises dentro da crise: respostas, incertezas e desencontros no combate à pandemia da Covid-19 no Brasil. Estud Av 2020; 34: 25–44.

11 Rezende LFM, Thome B, Schveitzer MC, et al. Adults at high-risk of severe coronavirus disease-2019 (Covid-19) in Brazil. Rev Saúde Pública 2020; 54. DOI:10.11606/s1518-8787.2020054002596.

12 de Souza WM, Buss LF, Candido D da S, et al. Epidemiological and clinical characteristics of the COVID-19 epidemic in Brazil. Nat Hum Behav 2020; 4: 856– 65.

13 Secretaria da saúde do estado de São Paulo. SIMI-SP: Pacientes internados por Síndrome Respiratória Aguda Grave (SRAG). https://www.saopaulo.sp.gov.br/planosp/simi/ (accessed Oct 25, 2020).

14 Niquini RP, Lana RM, Pacheco AG, et al. SRAG por COVID-19 no Brasil: descrição e comparação de características demográficas e comorbidades com SRAG por influenza e com a população geral. Cad Saúde Pública 2020; 36: e00149420.

15 Hasell J, Mathieu E, Beltekian D, et al. A cross-country database of COVID-19 testing. Sci Data 2020; 7: 345.

16 Telles E, Paschel T. Who Is Black, White, or Mixed Race? How Skin Color, Status, and Nation Shape Racial Classification in Latin America. Am J Sociol 2014; 120: 864–907.

17 IBGE - Instituto Brasileiro de Geografia e Estatística (2020). Pesquisa Nacional por Amostra de Domicílios Contínua (PNAD) COVID-19. Microdados..

18 Peixoto PS, Marcondes D, Peixoto C, Oliva SM. Modeling future spread of infections via mobile geolocation data and population dynamics. An application to COVID-19 in Brazil. PLOS ONE 2020; 15: e0235732.

19 De Souza Santos AA, Da Silva Cândido D, Marciel De Souza W, et al. SARS- CoV-2 non-pharmaceutical interventions in Brazilian municipalities. 2020; : 1046406 bytes.

20 Buss LF, Prete CA, Abrahim CMM, et al. Three-quarters attack rate of SARS- CoV-2 in the Brazilian Amazon during a largely unmitigated epidemic. Science 2020; published online Dec 8. DOI:10.1126/science.abe9728.

21 Weill JA, Stigler M, Deschenes O, Springborn MR. Social distancing responses to COVID-19 emergency declarations strongly differentiated by income. Proc Natl Acad Sci 2020; 117: 19658–60.

22 Chang S, Pierson E, Koh PW, et al. Mobility network models of COVID-19 explain inequities and inform reopening. Nature 2020; : 1–8.

23 Lustig N, Pabon VM, Sanz F, Younger SD. The Impact of COVID-19 Lockdowns and Expanded Social Assistance on Inequality, Poverty and Mobility in Argentina, Brazil, Colombia and Mexico. ECINEQ, Society for the Study of Economic Inequality, 2020 https://ideas.repec.org/p/inq/inqwps/ecineq2020-558.html (accessed Nov 20, 2020).

24 Giest S, Samuels A. ‘For good measure’: data gaps in a big data world. Policy Sci 2020; 53: 559–69.

25 Duranton G, Overman HG. Testing for Localization Using Micro-Geographic Data. Rev Econ Stud 2005; 72: 1077–106.

26 Souza PHGF de. A distribuição de renda nas pesquisas domiciliares brasileiras: harmonização e comparação entre Censos, PNADs e POFs. Rev Bras Estud Popul 2015; 32: 165–88.

27 Paim J, Travassos C, Almeida C, Bahia L, Macinko J. The Brazilian health system: history, advances, and challenges. The Lancet 2011; 377: 1778–97.

28 Nogueira Avelar e Silva R, Russo G, Matijasevich A, Scheffer M. Covid-19 in Brazil has exposed socio-economic inequalities and underfunding of its public health system. The BMJ. 2020; published online June 19. https://blogs.bmj.com/bmj/2020/06/19/covid-19-in-brazil-has-exposed-deeply-rooted-socio-economic-inequalities-and-chronic-underfunding-of-its-public-health-system/ (accessed Nov 20, 2020).

29 Castro MC, Carvalho LR de, Chin T, et al. Demand for hospitalization services for COVID-19 patients in Brazil. medRxiv 2020; : 2020.03.30.20047662.

30 Emanuel EJ, Persad G, Kern A, et al. An ethical framework for global vaccine allocation. Science 2020; 369: 1309–12.

