## Supplementary Appendix for "Social and racial inequalities in COVID-19 risk of hospitalisation and death across São Paulo state, Brazil"

Sabrina L Li, MSc [1*#], Rafael H M Pereira, DPhil [2*#], Carlos A Prete Jr, MSc [3*], Alexander E Zarebski, PhD [4], Lucas Emanuel, PhD [2], Pedro JH Alves, MSc [2], Pedro S Peixoto [5], PhD, Carlos K Braga, MSc [2], Andreza Aruska de Souza Santos, PhD [6], William M de Souza, PhD [4,7], Rogerio J Barbosa, PhD [8], Lewis F Buss, MD [9], Alfredo Mendrone-Junior, MD [10], Cesar de Almeida-Neto, MD [10,11], Suzete C Ferreira, PhD [10,12], Nanci A Salles, BSc [10,12], Izabel Marcilio, MD [13], Chieh-Hsi Wu, PhD [14], Nelson Gouveia, MD [15], Vitor Heloiz Nascimento, PhD [3], Ester C Sabino, MD [9], Nuno R Faria, PhD [4,9,16], Jane P Messina, PhD [1,17]

1. School of Geography and the Environment, University of Oxford, Oxford, United Kingdom.
2. Institute for Applied Economic Research, Brasília, Brazil.
3. Department of Electronic Systems Engineering, University of São Paulo, São Paulo, Brazil.
4. Department of Zoology, University of Oxford, Oxford, United Kingdom.
5. Department of Applied Mathematics, Institute of Mathematics and Statistics, University of São Paulo, São Paulo, Brazil.
6. Oxford School of Global and Area Studies, Latin American Centre, University of Oxford, Oxford, United Kingdom.
7. Virology Research Center, University of São Paulo, Ribeirão Preto, Brazil.
8. Center for Metropolitan Studies, Faculty of Philosophy, Languages and Literature, and Human Sciences, University of São Paulo, São Paulo, Brazil.
9. Departamento de Molestias Infecciosas e Parasitarias & Instituto de Medicina Tropical da Faculdade de Medicina da Universidade de São Paulo, São Paulo, Brazil.
10. Fundação Pró-Sangue Hemocentro de São Paulo
11. Disciplina de Ciências Médicas, Faculdade de Medicina da Universidade de São Paulo, São Paulo, Brazil.
12. Laboratory of Medical Investigation in Pathogenesis and Directed Therapy in Onco-Immuno – Hematology (LIM-31) HCFMUSP, University of São Paulo Medical School, São Paulo, Brazil.
13. Epidemiologic Surveillance Center, Hospital das Clinicas – University of São Paulo Medical School, São Paulo, Brazil.
14. Mathematical Sciences, University of Southampton, Southampton, United Kingdom.
15. Department of Preventive Medicine, University of São Paulo Medical School

16. MRC Centre for Global Infectious Disease Analysis, J-IDEA, Imperial College London, London, UK.
17. Oxford School of Global and Area Studies, University of Oxford, Oxford, United Kingdom.

*These authors contributed equally.

#Corresponding authors:

1. Sabrina L. Li, School of Geography and the Environment, University of Oxford, South Parks Rd, Oxford, OX1 3QY, United Kingdom.

2. Rafael H.M. Pereira, Institute for Applied Economic Research (Ipea), SBS - Quadra 1 - Bloco J - Ed. BNDES - CEP: 70076-900 - Brasília - DF – Brasil.

### Appendix A: Methodology details

#### 1 Probability of hospitalisation and death

Probability of hospitalisation and death due to COVID-19 were calculated by income quantile, race or administrative type of the health facility (public or private). The same method was used to calculate the estimates for the different grouping criteria (i.e., income quantile, race, administrative type of the health facility). We use the term 'class' to refer to a specific subgroup for each of these criteria, i.e., the higher income class, in the case of analysis by income, or the public health facility class in case of analysis by type of health facility. It is worth noting that only hospitalisations or deaths contained in the SIVEP-Gripe dataset are considered SARI cases. We also discarded cases that were confirmed to be the results of other etiological agents.

The health facility where each patient was hospitalized is defined for all SARI patients in the SIVEP-Gripe dataset. Administrative types of health facilities were extracted from the National Registry of Health Facilities (CNES) database. We classified as "Other" all health facilities whose administrative type is not marked as "SUS" (Brazilian public health system) or "Private" in the database, but health facilities with missing administrative type were discarded. In most cases, these are either public health facilities that accept patients with health insurance, or private health facilities that accept patients from the public health system. 26·7% of all SARI patients were notified by health facilities classified as public, compared to 30·2% for private, 21·5% for "Other" and 21·7% for health facilities with missing administrative type.

##### 1.1 Probability of death

Let $w_{ik}$ be the probability of the patient $i$ belonging to a given class $k$ in the grouping criterion of interest (income, race or administrative type of the health facility). When comparing private and public hospitals, $w_{ik}$ can only be 1 or 0 because the health facility type is known for all patients. Similarly, $w_{ik}$ can only be 1 or 0 for individuals whose races are not missing.

In the case of incomplete information, that is, individuals with unknown race and for the income comparisons, we assign a distribution over the possible values using estimates obtained from census data for each census tract. The probabilities $w_{ik}$ are assigned as the proportion of the individuals in the corresponding census tract that belong to class $k$ (that is, $w_{ik}$ is given by the racial or income distribution of the census tract where the individual lives). In order to compute the probabilities of death and SARI for a given class, the weights $w_{ik}$ are used to divide the contribution of each individual $i$ between all classes $k$ for $k = 1, \ldots, K$, where $K$ is the number of classes. Individuals whose census tract is not known were excluded from the income analysis, but only individuals with both unknown race and census tract were excluded from the racial analysis.

Let us denote the probability of an event $A$ as $P(A)$. The probability of death given class, age, and sex $P(\text{death}|\text{class}, \text{age}, \text{sex})$ was estimated by dividing the number of SARI deaths for a given class and age-sex group by the number of SARI cases for the same class, age, and sex group. The probability of death given class (among SARI patients) can be obtained through

$$P(\text{death}|\text{class}) = \sum_{\text{age,sex}} P(\text{death}|\text{class}, \text{age}, \text{sex}) P(\text{age}, \text{sex}|\text{class}),$$

where $P(\text{age}, \text{sex}|\text{class})$ is the conditional age-sex distribution of SARI patients given their class. This distribution may differ significantly for different classes due to differences in the age composition of classes, especially when race is used as class: The proportion of residents of the state of São Paulo that are older than 70 years old is 6·1% for White, 2·9% for *Pardo*, 4·4% for Black and 11·4% for Asians according to the 2010 census. Since the probability of death depends heavily on age, in order to allow a fairer comparison of the burden of the disease according to income and race, we use an age-sex normalised death probability given class, which is the death probability given class that would be obtained if all classes had the same age-sex distribution. If this age-sex normalisation was not applied, the probability of death would be overestimated for white patients and underestimated for black patients, as the white population is older than the black population. This normalised distribution is obtained by substituting $P(\text{age}, \text{sex}|\text{class})$ by $P(\text{age}, \text{sex})$, the proportion of patients from an age-sex group regardless of the class:

$$P_{\text{normalised}}(\text{death}|\text{class}) = \sum_{\text{age,sex}} P(\text{death}|\text{class}, \text{age}, \text{sex}) P(\text{age}, \text{sex})$$

This age-sex standardisation was not employed for the health facility type because patients from public and private hospitals share similar age-sex distributions. The age of the patients was discretized into the following groups: 0-9, 10-19, 20-29, 30-39, 40-49, 50-59, 60-69, 70-79, 80-89, 90+. It is worth noting that weighting estimates that depend on age to produce an age-standardised estimate was also done in other similar studies [1].

##### 1.2 Probability of hospitalisation by SARI

The probability of hospitalisation by severe acute respiratory infection (SARI) given each class for each criterion (only income or race in this analysis) is computed for each age-sex group as the ratio between the number of recorded SARI cases for a given class, age and sex and the number of individuals of that class and age-sex group. The age-sex distributions of for population of São Paulo was extracted from the 2010 census. The same age-sex standardisation employed to compute the probabilities of death was used to estimate the probabilities of SARI, but as in the 2010 census, patients above 70 years old were aggregated into the same age group. Only the race and income quantile classes are used in this analysis.

##### 1.3 Confidence intervals

Confidence intervals were estimated through bootstrapping using 1 000 realizations. For each realization, the same number of individuals in the dataset were randomly selected with replacement from the set of individuals in the dataset. Then, the desired estimates and odds ratios are obtained from each resampled dataset. For the racial analysis each selected individual $i$ with unknown race had its race imputed for each bootstrap iteration such that the probability of the imputed race being $k$ is $w_{ik}$. The confidence intervals were obtained from the quantiles of the set of 1 000 bootstrapped parameters.

#### 2 Seroprevalence by socioeconomic status

The broader risk of infection of SARS-CoV-2 beyond hospitalisation was analysed using seroprevalence data from blood donors collected in the city of São Paulo by [2]. Given that samples were taken across the city, a population-weighted cluster sample of approximately 1 000 blood donations were tested each month between February and August 2020 using a chemiluminescence assay that detects IgG against the SARS-CoV-2 nucleocapsid (N) protein (Abbott, Chicago, USA). Self-reported race and education level were recorded at the time of blood donation. To correct for differences in the age-sex distribution of blood donors compared to the population of São Paulo, we applied an age-sex normalisation to the measured prevalence. Details about the data collection methods can be found in [3]. We calculated the proportion of individuals by education and race category with detectable anti-SARS-CoV-2 antibodies. 95% confidence intervals were calculated by the exact binomial method and corrected for the specificity and sensitivity of the test [4].

#### 3 Geospatial analysis

##### 3.1 Model description

We used a Bayesian hierarchical model to compute the relative risk of hospitalisation at the municipality level for São Paulo state (n= 645) and at the census tract level for the greater metropolitan area of São Paulo (GMSP: n=30 815). The number of observed cases $Y_i$ in an area $i$ is modelled using a Poisson distribution $Y_i \sim Poisson(\lambda_i)$ with mean $\lambda_i = E_i\ \mu_i$ where $E_i$ is the expected number of cases in area $i$ under a null model in which cases are uniformly distributed among the population, i.e., the number of cases in a given area is proportional to the population of that area. For each area $i$, this is given by $E_i = \frac{\sum_i Y_i}{\sum_i pop_i} \times\ pop_i$, where $pop_i$ is the population in area $i$. The factor of $\mu_i$ describes the area-specific risk and models the additional variation in the observation process [5].

To quantify the uncertainty in the point estimates of the mean relative risk estimates, we mapped the posterior probability of elevated relative risk in each area (Appendix B - Figure S6). This is the posterior probability that a tract has an elevated risk of observing cases, formally P($\mu_i$ > 1| data). For instance, a posterior probability of 0.6 in an area indicates a 60% chance that this area is at greater risk of observing cases.

##### 3.2 Modelling relative risk

We fit a log-linear model to estimate the relative risk $\mu_i$, which is modelled as the sum of an intercept and random effects. Random effects are broken into the spatial ($A_i$) and temporal components ($B_i$), as shown in Eq. (1·1):

$$\log(\mu_i) = \alpha + A_i + B_i \quad (1\cdot1)$$

$$\log(\mu_i) = \alpha + U_i + V_i + \gamma_t + \phi_t \quad (1\cdot2)$$

To account for existing spatial autocorrelation, we used a Besag-York-Mollié model (BYM) [6] to separate the spatial component into spatially structured $U_i$, and non-spatial, unstructured random effects, $V_i$, so ($A_i = U_i + V_i$), as shown in Eq. 1·2. In the BYM model, a conditional autoregressive (CAR) process is used to introduce correlation among the $U_i$ for each tract. Given the $U_i$ of neighbouring areas, the $U_i$ has a normal distribution with mean equal to the average of the neighbours' $U_i$, and variance $s_i^2 = \frac{1}{\#N(i)\tau_U}$ where $\#N(i)$ is the number of areas that share boundaries with area $i$ and $\tau_U$ is a precision parameter. The random effect, $V_i$ follows a zero mean normal distribution with precision parameter, $\tau_V = \frac{1}{\sigma_{v^2}}$ (where $\sigma_{V^2}$ is the variance). Both random effects in the model capture extra-Poisson variability, and were expressed as the following:

$$U_i |\ U_{j \neq i} \sim \mathcal{N}\left(m_i, s_i^2\right), \quad V_i \sim \mathcal{N}(0, \sigma_v^2)$$

$$m_i = \frac{\sum_{j \in N(i)} U_j}{\# N(i)} \quad , \quad s_i^2 = \frac{\sigma_U^2}{\# N(i)} = \frac{1}{\# N(i) \tau_U}$$

To account for temporal structure in the data, we included the random effect ($B_t = \gamma_t + \phi_t$), which assumes that the number of cases observed in a given area depends on the number of cases observed in the given area in the previous month and a residual [5,7]. The temporal component includes $\gamma_t$, a temporally structured effect modelled dynamically using a random walk of order 1, and an unstructured temporal effect $\phi_t$ to account for independent time effects, which follows a zero mean normal distribution. Both are expressed as the following:

$$\gamma_t | \gamma_{t-1} \sim \mathcal{N}\left(\gamma_{t-1}, \frac{1}{\tau_\gamma}\right)$$

$$\phi_t \sim \mathcal{N}\left(0, \frac{1}{\tau_\phi}\right)$$

We adopted minimally informative prior distributions in R-INLA [3]. The log of the precision parameters adopted for the spatial effects, $\tau_U$ and $\tau_V$ , follows a gamma distribution with shape 1 and rate 0·0005. The precision parameter for both the structured and unstructured temporal effects $\tau_\gamma$ and $\tau_\phi$ also follows a gamma distribution, with shape 1 and rate 0·001. The prior default distributions in R-INLA, which are the recommended settings [4], were also used for the precision parameters of both $U_i$, $V_i$, $\gamma_t$, and $\phi_t$.

##### 3.3 Ecological regression

To evaluate the effects of socioeconomic covariates on the risk of hospitalisation at the municipality level, we reformulated our model expressed by Eq. 1·3 by adding a fixed effect, which we refer as the ecological regression model. The log of the relative risk is given by the following:

$$\log(\mu_i) = \alpha + U_i + V_i + \gamma_t + \phi_t + X'_{ik}\, \beta \ (1{\cdot}3)$$

where $X'_{ik}$ is the $i$ th row and $k$th column of covariates matrix $X$ based on the socioeconomic covariates for each municipality and month, and $\beta$ is the regression parameter modelled as fixed effects with normal priors ($\beta \sim \mathcal{N}(0, 100)$ ) .

Here, we define $U_i$ using a graphical structure in R-INLA which describes the connections between municipalities by looking at estimates of the level of human mobility between them in the state of São Paulo. We considered two municipalities to be connected if there were at least 550 journeys between them. This threshold was selected to ensure that the sparsity of the connectivity matrix was similar to the nearest neighbour matrix described in Section 3·2. The number of origin-destination journeys between municipalities are retrieved from processed mobile geo-location data obtained from the In Loco company described elsewhere [8]. The prior default distributions in R-INLA were used for the precision parameters of both $U_i$, $V_i$, $\gamma_t$, and $\phi_t$.

##### 3.4 Computational method

We carried out model fitting with R-INLA which uses an Iterated Nested Laplace Approximation (INLA) based on a combination of analytical approximations and numerical integration to estimate posterior distributions [9]. INLA was designed as an efficient alternative to Markov Chain Monte Carlo (MCMC), which is both computationally and time-intensive when applied to a large amount of data. It can be suitably applied to latent Gaussian models including generalised linear models to spatial and spatio-temporal models.

##### 3.5 Model evaluation

We evaluated our model by plotting the empirical relative risk in each area against the fitted risk determined by our model (Figure A1). The empirical relative risk was calculated by weighting the total number of observed cases in a given municipality, with municipality level population as a proportion of the entire state of São Paulo (defined as offset). A density plot illustrating the distribution of empirical versus predicted risk was also created to assess model fit (Figure A2).

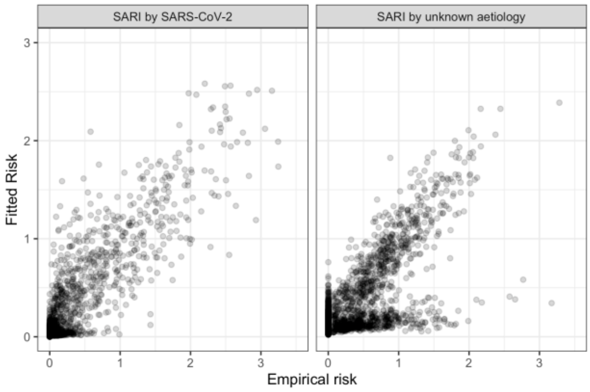

Figure A1. Empirical vs. fitted risk

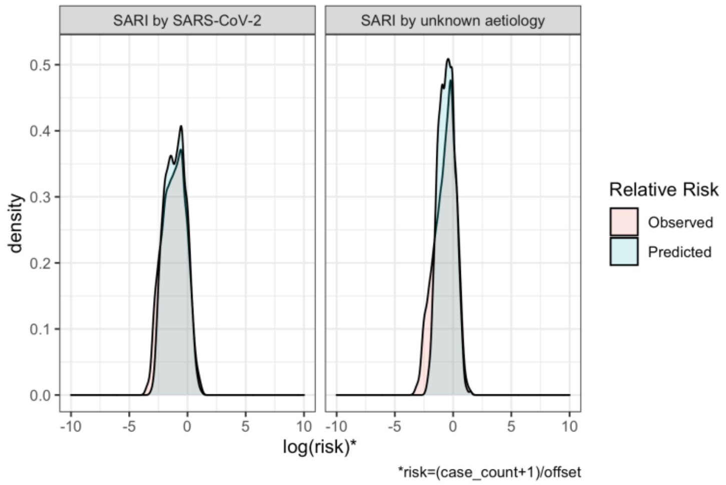

Figure A2. The estimated value of the log-risk for each area closely matches the observed risk in each area.

#### 3.6 Covariates

##### 3.6.1 2010 census

Data on socioeconomic covariates used in the ecological regression at the municipality level were obtained from the 2010 population census compiled by the Brazilian Institute of Geography and Statistics (IBGE) [10].These data include the average household income per capita, housing density (number of private households/km$^2$), sanitation conditions (proportion of population with access to piped water, access to sewage network, and/or access to septic tank), percentage of urban and rural population, and income inequality measured with the Gini index.

To our knowledge, this is the most complete and recent socioeconomic dataset available for Brazil at our required spatial resolution since no census was carried out in 2020. To assess the variability of some of these covariates over a ten year period, we obtained the same socioeconomic data from the 2000 IBGE census. We compared the same covariates by plotting them at the municipality level for a small sample (in this case, we decided to extract the 39 municipalities in RMSP) (Figure A3). We observed a general trend of increase in income per capita, household density, and number of residents per household, at the municipality level, between 2000 and 2010. Given the inconsistent variation in population access to piped water, septic tank, and sewage network by municipality over the 10-year time frame, we decided to omit these variables from the analysis.

To improve accuracy, we decided to compute population density (people/km$^2$) at the census tract level, then aggregating it for each municipality, instead of using household density. This was calculated by dividing the total resident population by the area of each census tract using data retrieved from the 2010 census. For large census tracts (with area greater than 0·12 km$^2$), we considered the occupied area rather than the total area of the census tract. This was done to improve the accuracy of population density estimates in large census tracts, particularly for those located in rural areas, by focusing only on human habitats. Human occupied areas were identified based on population counts from a fine regular grid of 200 meters, which was generated by IBGE for the 2010 census using fieldwork and satellite imagery data.

We checked for multicollinearity by assessing the correlation between variables and computing the variation inflation factor (VIF) of each covariate. Based on these tests, we removed the proportion of informal workers and unemployment from our final model to avoid multicollinearity with the variables on household density and income per capita.

##### 3.6.2 Distance to the nearest health facility

We have also computed mean distance to the nearest health facility of each municipality. To do this, we calculated the road network distance from the centroid of each census tract to the nearest healthcare facility in R with the dodgr package [11]. We considered all the 830 healthcare facilities registered in the SIVE-Gripe database in São Paulo state that hospitalised SARI patients via the Unified Health System (SUS).

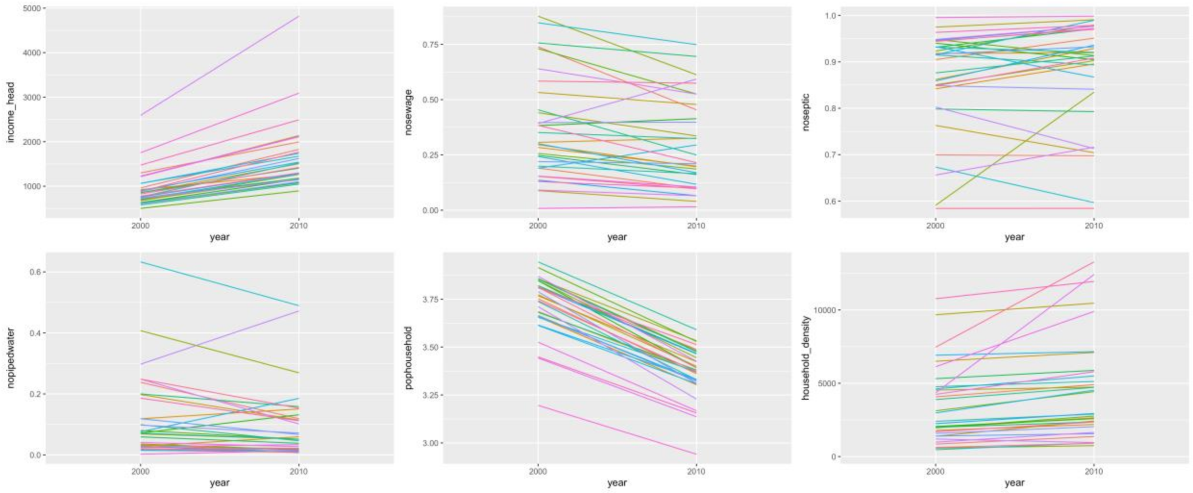

Figure A3. Trends in covariate values between 2000 and 2010. Each coloured line represents a municipality within the GMSP (n= 39).

##### 3.7 Spatial joins for the GMSP

To improve the accuracy of modelling small areas for the RMSP, we performed spatial joins of census tracts to ensure that each census tract had at least one SARI case. Spatial joins were performed by selecting the census tract with the smallest population and joining it to its nearest neighbour (determined by the shortest distance between centroids). This was repeated until all tracts had at least one case of SARI.

#### 4 Probability of working conditions

We used PNAD COVID-19 data from May to September 2020. During this period, 1 888 560 individuals were interviewed, of whom 171 480 were living in São Paulo state. For each individual, the PNAD COVID-19 survey also collects self-reported data on access to COVID-19 testing and comorbidities. From July, August, and September 2020, interviewees were also asked about comorbidities.

In the PNAD COVID-19 survey, work status is a categorical variable that can assume four values: face-to-face, telework, paid leave, and unpaid leave. We model the conditional probabilities using a multinomial logistic regression.

$$\mathrm{P}(y_i = m|\boldsymbol{x}_i) = \frac{\exp(\boldsymbol{x}_i\boldsymbol{\beta}_m)}{\sum_{j=1}^{4}\exp(\boldsymbol{x}_i\boldsymbol{\beta}_j)}$$

$\mathrm{P}(y_i = m|\boldsymbol{x}_i)$ represents the probability that, for the individual $i$, the variable Work Status ($y$) will assume a particular value $m$, given a $k \times 1$ vector $\boldsymbol{x}_i$ of explanatory variables and an intercept. The letter $j$ varies from 1 to 4 and indexes the four categories of Work Status. $\boldsymbol{\beta}_j$ is a vector of coefficients for the category $j$. The reference category ($j = 1$) is Face-to-Face work and, by construction, $\boldsymbol{\beta}_1 = \boldsymbol{0}$. So, the estimated vector of parameters $\widehat{\boldsymbol{\beta}}$ is $3k \times 1$ dimensional:

$$\hat{\boldsymbol{\beta}} = \begin{bmatrix} \hat{\boldsymbol{\beta}}_T \\ \hat{\boldsymbol{\beta}}_P \\ \hat{\boldsymbol{\beta}}_U \end{bmatrix}$$

Where $\underbrace{\hat{\boldsymbol{\beta}}_{\mathrm{T}}}_{k\times 1} = \hat{\boldsymbol{\beta}}_{\mathrm{Telework|Face\text{-}to\text{-}face}}$, $\underbrace{\hat{\boldsymbol{\beta}}_{\mathrm{P}}}_{k\times 1} = \hat{\boldsymbol{\beta}}_{\mathrm{Paid\ leave|Face\text{-}to\text{-}face}}$, and $\underbrace{\hat{\boldsymbol{\beta}}_{\mathrm{U}}}_{k\times 1} = \hat{\beta}_{\mathrm{Unpaid\ leave|Face\text{-}to\text{-}face}}$.

We estimated four different nested models, by adding explanatory variables stepwise. Model 0 has race, sex (a dummy variable) and age (a 3rd degree orthogonal polynomial). Model 1 adds education, Model 2 adds occupation (ISCO-08 1-digit groups – see Table **A2**) and a dummy variable for (in)formality, Model 3 adds dummy variables for the months the observations were collected in PNAD COVID-19.

In order to calculate predicted probabilities, we let the variable of interest (e.g., race) to vary and set all other explanatory variables to their grand mean. This way, all the probability differences between categories (e.g. White, Black, *Pardo*, and Asian) are due to changes in the variable of interest.

Models were estimated taking into account the Complex Sample Design of PNAD COVID-19. So, the Variance-Covariance Matrix of the Coefficients already take into account heteroskedasticity and autocorrelation between observations. Confidence intervals for the predicted probabilities were calculated by parametric bootstrapping. By the Central Limit Theorem, the estimated coefficients follow a Multivariate Normal Distribution:

$$\hat{\boldsymbol{\beta}} \sim \mathcal{N}(\boldsymbol{\beta}, \boldsymbol{\Sigma}_\beta)$$

Using $\hat{\boldsymbol{\beta}}$ as the mean and $\underbrace{\hat{\boldsymbol{\Sigma}}_\beta}_{3k\times 3k}$ as the variance-covariance matrix, we simulated 2 000 samples of coefficients. We organized the simulated coefficients for each category of dependent variable in separate matrices: $\underbrace{\hat{\mathbf{B}}_{\mathrm{T}}^{sim}}_{2000\times k}$, $\underbrace{\hat{\mathbf{B}}_{\mathrm{P}}^{sim}}_{2000\times k}$, and $\underbrace{\hat{\mathbf{B}}_{\mathrm{U}}^{sim}}_{2000\times k}$. For a given vector of observations $\boldsymbol{x}$, a distribution of fitted values can be calculated as:

$$\underbrace{\boldsymbol{x}}_{1\times k} \underbrace{\left(\hat{\boldsymbol{\beta}}_{\mathrm{T}}^{sim}\right)^T}_{k\times 2000} = \underbrace{\hat{\boldsymbol{\mu}}_{\mathrm{T}}^{sim}}_{1\times 2000}$$
$$\underbrace{\boldsymbol{x}}_{1\times k} \underbrace{\left(\hat{\boldsymbol{\beta}}_{\mathrm{P}}^{sim}\right)^T}_{k\times 2000} = \underbrace{\hat{\boldsymbol{\mu}}_{\mathrm{P}}^{sim}}_{1\times 2000}$$
$$\underbrace{\boldsymbol{x}}_{1\times k} \underbrace{\left(\hat{\boldsymbol{\beta}}_{\mathrm{U}}^{sim}\right)^T}_{k\times 2000} = \underbrace{\hat{\boldsymbol{\mu}}_{\mathrm{U}}^{sim}}_{1\times 2000}$$

And, by definition $\underbrace{\hat{\boldsymbol{\mu}}_{\mathrm{F}}^{sim}}_{1\times 2000} = \underbrace{\mathbf{0}}_{1\times 2000}$. A distribution of predicted probabilities given $\boldsymbol{x}$ is given by:

$$\underbrace{\mathbf{P}(y_i = m|\boldsymbol{x}_i)}_{1\times 2000} = \frac{\exp\left(\hat{\boldsymbol{\mu}}_m^{sim}\right)}{\sum_{j=1}^{4} \exp\left(\hat{\boldsymbol{\mu}}_j^{sim}\right)}$$

A 95% confidence interval is obtained if we take the quantiles 0,025 and 0,975 of this vector of predicted probabilities.

#### 5 Event-study analysis

We used an event study model to investigate how people from different socioeconomic groups changed their daily isolation levels after the implementation of state non-pharmaceutical intervention (NPI). The model was conducted using an ecological analysis where both socioeconomic characteristics of the population and daily isolation levels are spatially aggregated on the H3 hexagonal grid at resolution 8. Each cell has an edge of approximately 460 meters and an area of 0·74 $km^2$. Hexagonal H3 cells were then ranked by income based on quintiles of average income per capita. Cells were also categorised as predominantly White when at least 60% of the population self-declared White and predominantly Black when at least 60% of the population self-declared Black or *Pardo*.

The racial composition and income level of each cell were determined using dasymetric interpolation of the 2010 census tract data in two steps. First, data on income and race were passed to a finer regular grid of 200 meters and linked with population count by finding the aerial intersection and population size of each cell. This was reaggregated from the regular grid to the hexagonal grid. Hexagonal H3 cells were then ranked by income based on quintiles of average income per capita. Cells were categorised as predominantly Black when at least 60% of the population self-declared Black or *Pardo*, and likewise for White.

In the event study model, our treated group is composed of hexagons predominantly of White population (race analysis) and the 20% wealthiest hexagons (income analysis). Conversely, the comparison groups were composed of hexagons with predominantly Black population and the 20% poorest hexagons. Our specification includes indicators for pre-and post-treatment effects, as follows:

$$Y_{id} = \left[ \sum_{\tau=-12}^{-2} \beta_\tau I(t_{id} - t^* = \tau) + \sum_{\tau=0}^{151} \beta_\tau I(t_{id} - t^* = \tau) \right] + X'_{id}\Theta + \omega_d + \mu_i + \varepsilon_{id}$$

where $Y_{id}$ is the outcome (daily isolation level) observed for hexagon $i$ at day $d$; the indicator $I(t_{id} - t^* = \tau)$ measures the time (in days) relative to the day of the state NPI implementation on date $t^*$. We set the coefficient $\beta_{-1}$ (March 12) equal to zero to use the day immediately prior to the state NPI implementation as the reference. $X'_{id}$ represents the set of hexagons covariates: a dummy variable indicating the beginning of NPI flexibilization period in each municipality, and a time-varying variable with the number of days relative to the first confirmed case of SARI in each hexagon (equates to 0 for days prior to the first case). $\mu_i$ is a hexagon fixed effect that controls non-parametrically for time-invariant hexagon factors, such as hexagons' fixed geographical aspects (e.g., urban infrastructure, proximity to healthcare facilities, urban density etc), $\omega_d$ is a day fixed effect that controls non-parametrically for aggregate shocks and other policies common to all hexagons at a specific moment in time, and $\varepsilon_{id}$ is an idiosyncratic error term. All observations are weighted by the size of resident population in each hexagon [10]. Finally, we clustered standard errors at the hexagon level to make estimations robust to serial correlation and heteroskedasticity [12].

This method allows us to more formally test for pre-trends in outcome variables in the pre-period. The identifying assumption is that the time trend in the mobility level in treated areas would have a similar trend as the one observed in similar nontreated areas in the absence of the policy intervention. Coefficient estimates of $\beta_\tau$; with $\tau < 0$ (representing the change, in percentage points, in the outcome each day pre-intervention) serves as a direct test of the plausibility of the identifying assumption. If hexagons have similar trends before the date of state declaration and diverge only after policy, it provides strong evidence that such changes were caused by the state NPI adoption rather than an unobservable factor.

#### 5.1 Summary statistics

Table 1 presents summary statistics for the main variables used in our analysis. Columns (1)-(3) shows the number of observations, the mean and the standard deviation for the treated group, while columns (4)-(6) exhibit the same statistics for the comparison group. Panel A shows the summary statistics for the race estimation sample, while Panel B presents the summary statistics for the income estimation sample.

Table A1 - Summary Statistics of hexagon characteristics

| | (1) | (2) | (3) | (4) | (5) | (6) |
|---|---|---|---|---|---|---|
| | Panel A – Race sample estimation | | | | | |
| | Black | | | White | | |
| | Obs. | Mean | Std. Dev | Obs. | Mean | Std. Dev |
| Daily isolation level | 16 187 | 0·418 | 0·084 | 225 231 | 0·457 | 0·100 |
| Days after first SARI case | 16 187 | 45·47 | 45·23 | 225 231 | 59·16 | 47·14 |
| Days with NPI flexibilization | 16 187 | 0·397 | 0·489 | 225 231 | 0·387 | 0·487 |
| | Panel B – Income sample estimation | | | | | |
| | Low Income | | | High Income | | |
| | Obs. | Mean | Std. Dev | Obs. | Mean | Std. Dev |
| Daily isolation level | 97 139 | 0·419 | 0·088 | 90 383 | 0·486 | 0·103 |
| Days after first SARI case | 97 139 | 47·55 | 45·81 | 90 383 | 63·40 | 47·44 |
| Days with NPI flexibilization | 97 139 | 0·396 | 0·489 | 90 383 | 0·381 | 0·486 |

Notes. This table polls all days of data per group in each sample estimation (from March 1 to August 11). Data are at the hexagon-by-day level.

#### 5.2 Model Sensitivity analysis

We tested how the results of the event study by running the regression with and without covariates and testing for different number of days after the introduction of NPIs. Table A2 shows the results including only one treatment variable so it represents the effect of the average treatment every day. With 99% of statistical significance in all scenarios, the results indicate that the effect tends to increase up until the 120$^{th}$ day. The effects of NPI on isolation levels is consistently positive and significant in all estimates. The inclusion of covariates does not significantly change the coefficients nor the confidence intervals.

Table A2 – Sensitivity analysis of NPI effects on isolation levels

| Number of days after NPI | Analysis by income | | | | | |
|---|---|---|---|---|---|---|
| | Without covariates | | | With covariates | | |
| | Mean | Min 95 | Max 95 | Mean | Min 95 | Max 95 |
| 10 | 0·055*** | 0·053 | 0·058 | 0·054*** | 0·052 | 0·057 |
| 30 | 0·084*** | 0·082 | 0·088 | 0·081*** | 0·079 | 0·085 |
| 90 | 0·087*** | 0·084 | 0·091 | 0·084*** | 0·081 | 0·088 |
| 120 | 0·083*** | 0·080 | 0·087 | 0·082*** | 0·079 | 0·086 |
| 150 | 0·079*** | 0·076 | 0·083 | 0·078*** | 0·075 | 0·082 |

| Number of days after NPI | Analysis by race | | | | | |
|---|---|---|---|---|---|---|
| | Without covariates | | | With covariates | | |
| | Mean | Min 95 | Max 95 | Mean | Min 95 | Max 95 |
| 10 | 0·039*** | 0·036 | 0·044 | 0·038*** | 0·035 | 0·043 |
| 30 | 0·064*** | 0·060 | 0·069 | 0·061*** | 0·057 | 0·066 |
| 90 | 0·062*** | 0·058 | 0·068 | 0·059*** | 0·055 | 0·064 |
| 120 | 0·059*** | 0·055 | 0·064 | 0·056*** | 0·053 | 0·061 |
| 150 | 0·055*** | 0·051 | 0·060 | 0·053*** | 0·049 | 0·058 |

Note. *** p <0·01

#### 6 Crosswalk: PNAD COVID-19 Occupations to ISCO-08 (1-digit)

The crosswalk from PNAD COVID-19 occupational codes to ISCO-08 1-digit codes was done in two steps. First, we applied the conversion rule presented in Table A1 to the occupational codes of occupation categories in PNAD COVID-19 (variable C007C). Then we classified the values 2 and 3 of variable C007 as "Military" (once these occupations are not registered in C007). We further disaggregated Health Professionals and Technicians into separate occupational categories.

**Table A2 – Crosswalk: Variable C007C to ISCO-08 1-digit groups**

| PNAD-COVID-19 Occ. Code | PNAD COVID-19 Occupations (English Label) | ISCO-08 (1-digit) | ISCO-08 Label |
|---|---|---|---|
| 1 | Domestic worker, daily cleaner, cook (in private households), | 9 | Elementary Occupations |
| 2 | Janitor, cleaning assistant, etc. (in public or private company), | 9 | Elementary Occupations |
| 3 | Office clerk | 4 | Clerical Support Workers |
| 4 | Secretary, receptionist | 4 | Clerical Support Workers |
| 5 | Telemarketing operator | 4 | Clerical Support Workers |
| 6 | Merchant (owner of bars or shops etc.) | 5 | Services and Sales Workers |
| 7 | Store salesperson | 5 | Services and Sales Workers |
| 8 | Home seller, sales representative, catalog seller | 5 | Services and Sales Workers |
| 9 | Street vendors | 9 | Elementary Occupations |
| 10 | Cook and waiter (for restaurants, companies) | 5 | Services and Sales Workers |
| 11 | Baker, butcher and confectioner | 5 | Services and Sales Workers |
| 12 | Farmer, animal breeder, fisherman, forester and gardener | 6 | Skilled Agricultural, Forestry and Fishery Workers |
| 13 | Agricultural labourers | 9 | Elementary Occupations |
| 14 | Drivers (ride hailing apps, taxi, van, mototaxi, bus) | 8 | Plant and Machine Operators and Assemblers |
| 15 | Truck driver | 8 | Plant and Machine Operators and Assemblers |

| | | | |
|---|---|---|---|
| 16 | Courier services by motorcycle | 9 | Elementary Occupations |
| 17 | Delivery of goods (restaurant, pharmacy, store, Uber Eats, IFood, Rappy etc.) | 9 | Elementary Occupations |
| 18 | Bricklayer, stonemasons, painter, electrician, carpenter | 7 | Craft and Related Trades Workers |
| 19 | Mechanic of vehicles, industrial machineries etc. | 7 | Craft and Related Trades Workers |
| 20 | Craftsman, dressmaker and shoemaker | 7 | Craft and Related Trades Workers |
| 21 | Hairdresser, manicure and other beauty occupations | 5 | Services and Sales Workers |
| 22 | Machine operator, assembler in the industry | 8 | Plant and Machine Operators and Assemblers |
| 23 | Production assistant, loading and unloading | 8 | Plant and Machine Operators and Assemblers |
| 24 | Teachers and professors (kindergarten, elementary, high school or higher education) | 2 | Professionals |
| 25 | Pedagogue, teacher of languages, music, art and tutoring | 2 | Professionals |
| 26 | Health professionals | 2 | Professionals |
| 27 | Health Technician | 3 | Technicians and Associate Professionals |
| 28 | Babysitters and personal caretakers | 5 | Services and Sales Workers |
| 29 | Security, vigilant, other guard security services | 5 | Services and Sales Workers |
| 30 | Civil police | 5 | Services and Sales Workers |
| 31 | Doorman or Porter | 9 | Elementary Occupations |
| 32 | Artist, religious (priest, pastor, etc.) | 2 | Professionals |
| 33 | Director, manager, political or commissioned position | 1 | Managers |
| 34 | Other higher-level profession (lawyer, engineer, accountant, journalist, etc.) | 2 | Professionals |
| 35 | Other mid-level technician or professional | 3 | Technicians and Associate Professionals |
| 36 | Others | 10 | OTHER |

### 7 References

1. Dowd JB, Andriano L, Brazel DM, Rotondi V, Block P, Ding X, et al. Demographic science aids in understanding the spread and fatality rates of COVID-19. Proc Natl Acad Sci. 2020;117:9696–8.

2. Buss LF, Prete CA, Abrahim CMM, Mendrone A, Salomon T, Almeida-Neto C de, et al. Three-quarters attack rate of SARS-CoV-2 in the Brazilian Amazon during a largely unmitigated epidemic. Science [Internet]. 2020 [cited 2020 Dec 8]; Available from: https://science.sciencemag.org/content/early/2020/12/07/science.abe9728

3. Buss LF, Prete CA, Abrahim CM, Mendrone A, Salomon T, Almeida-Neto C de, et al. COVID-19 herd immunity in the Brazilian Amazon. medRxiv. 2020;2020.09.16.20194787.

4. Diggle PJ. Estimating Prevalence Using an Imperfect Test [Internet]. Vol. 2011, Epidemiology Research International. Hindawi; 2011 [cited 2020 Nov 20]. p. e608719. Available from: https://www.hindawi.com/journals/eri/2011/608719/

5. Lawson AB. Bayesian Disease Mapping: Hierarchical Modeling in Spatial Epidemiology, Second Edition. CRC Press; 2013. 398 p.

6. Besag J, York J, Mollié A. Bayesian image restoration, with two applications in spatial statistics. Ann Inst Stat Math. 1991;43:1–20.

7. Knorr-Held L. Bayesian modelling of inseparable space-time variation in disease risk. Stat Med. 2000;19:2555–67.

8. Peixoto PS, Marcondes D, Peixoto C, Oliva SM. Modeling future spread of infections via mobile geolocation data and population dynamics. An application to COVID-19 in Brazil. PLOS ONE. 2020;15:e0235732.

9. Rue H, Martino S, Chopin N. Approximate Bayesian inference for latent Gaussian models by using integrated nested Laplace approximations. J R Stat Soc Ser B Stat Methodol. 2009;71:319–92.

10. Censo Demográfico | IBGE [Internet]. [cited 2020 Nov 20]. Available from: https://www.ibge.gov.br/estatisticas/sociais/populacao/9662-censo-demografico-2010.html?=&t=o-que-e

11. Padgham M. dodgr: An R Package for Network Flow Aggregation. Findings. 2019;6945.

12. Bertrand M, Duflo E, Mullainathan S. How Much Should We Trust Differences-In-Differences Estimates? Q J Econ. 2004;119:249–75.

#### Appendix B: Figures

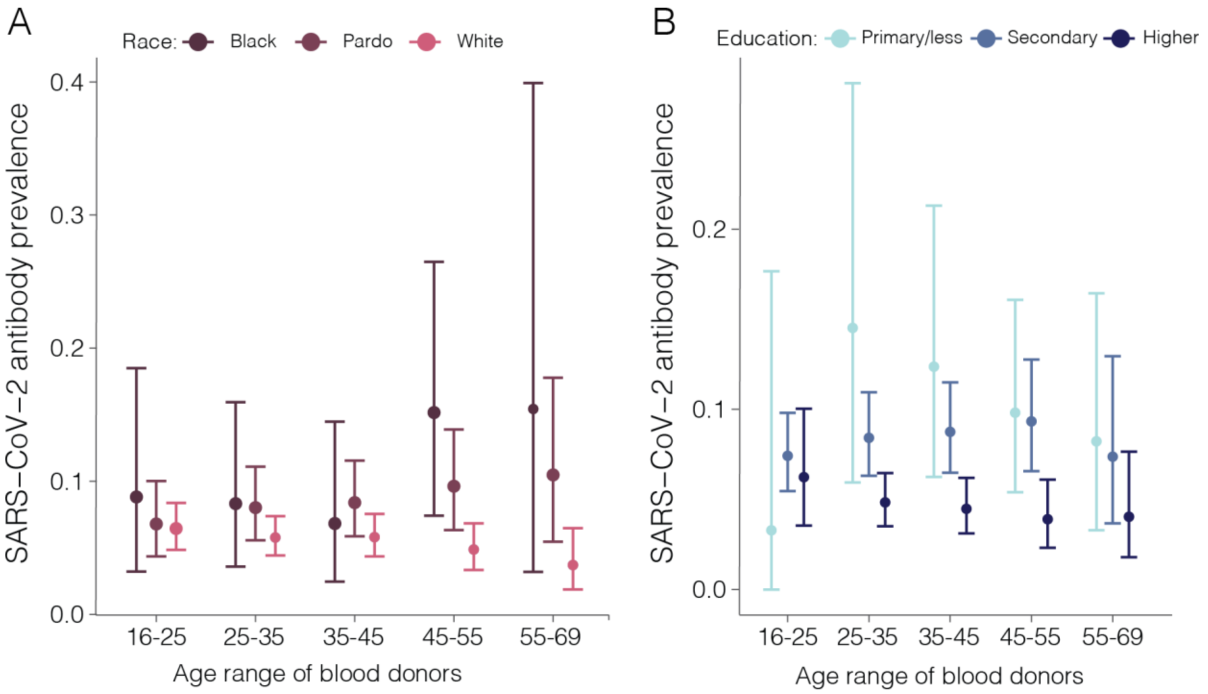

Figure S 1. Prevalence of anti-SARS-CoV-2 IgG antibodies among blood donors in São Paulo, Brazil, according to self-reported race and education attainment. Source: Covid-IgG study [2].

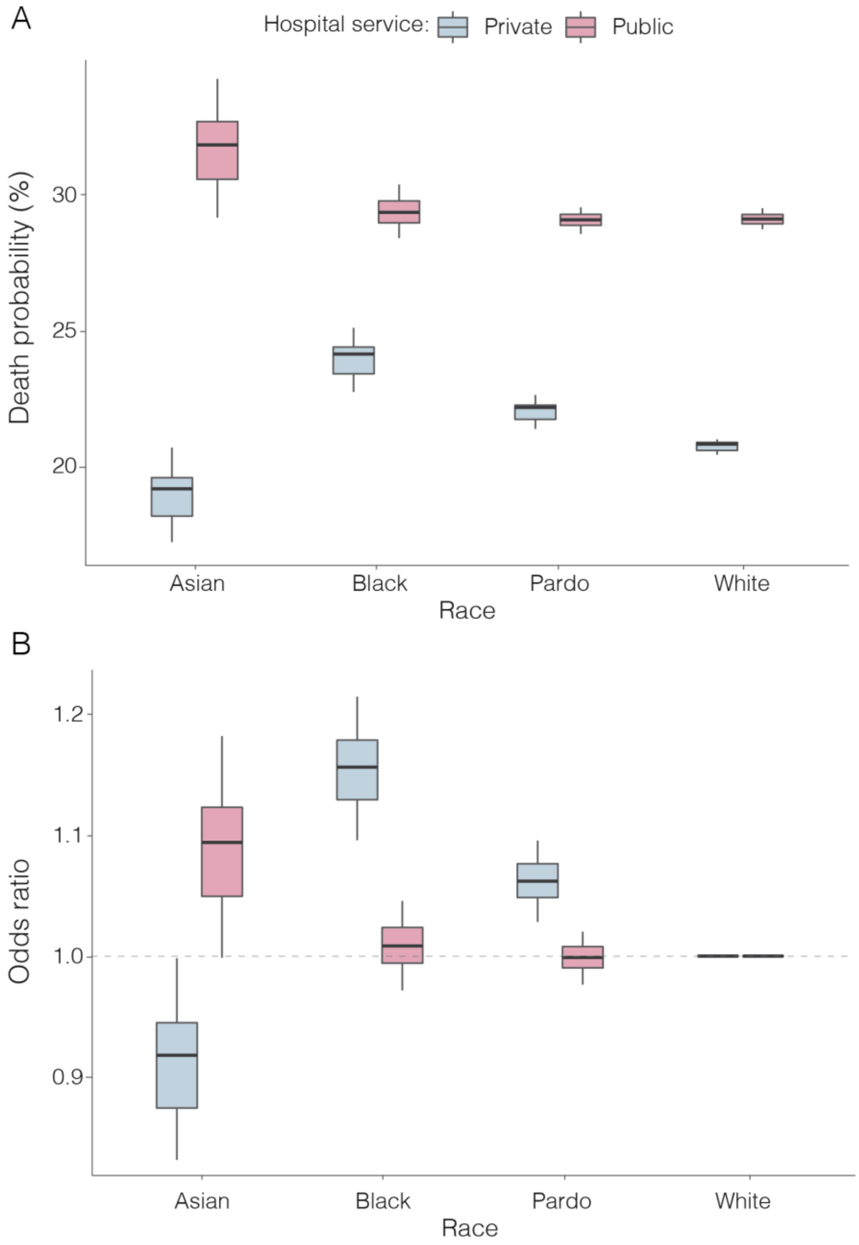

Figure S 2. Death probability and odds ratio of death by race in each hospital type. Source: SIMI-SP.

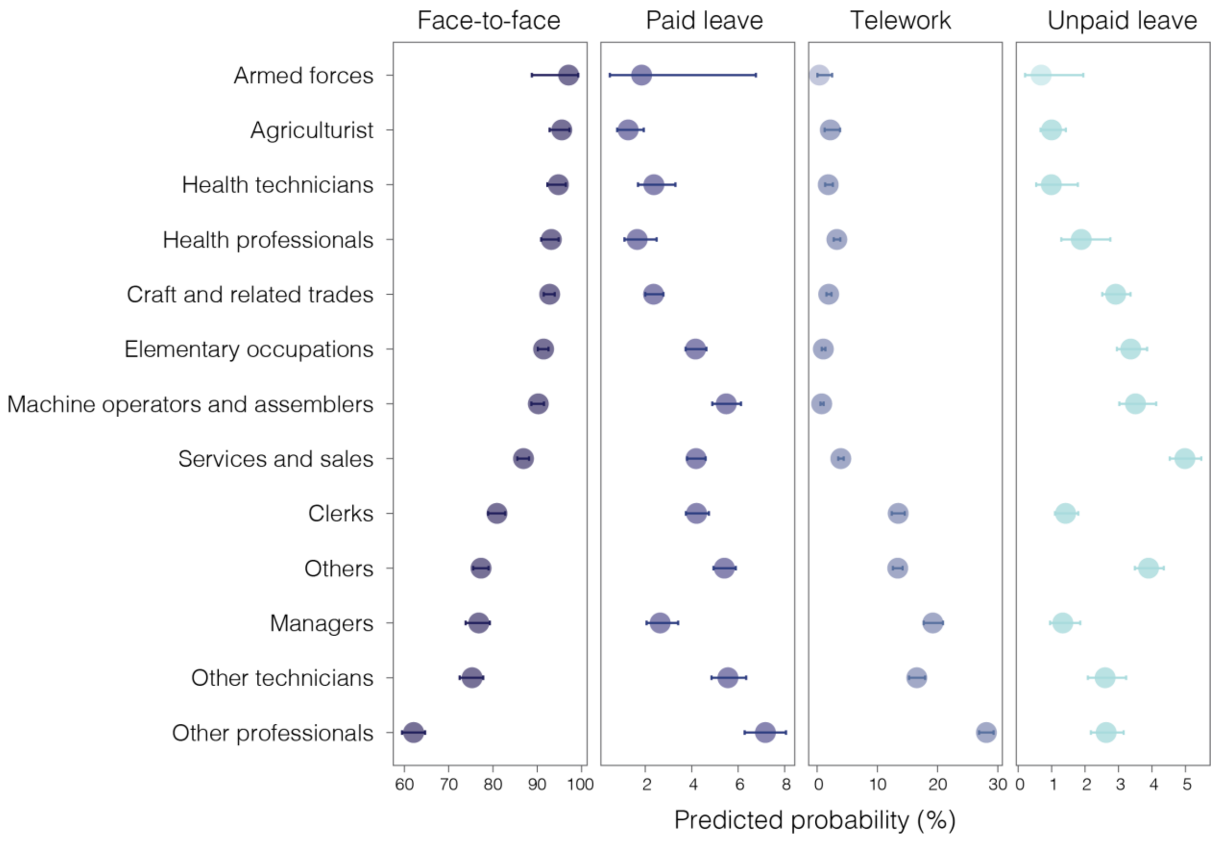

Figure S 3. Probability of working condition by occupation type between May and September 2020 in São Paulo state. Source: PNAD COVID-19 (IBGE).

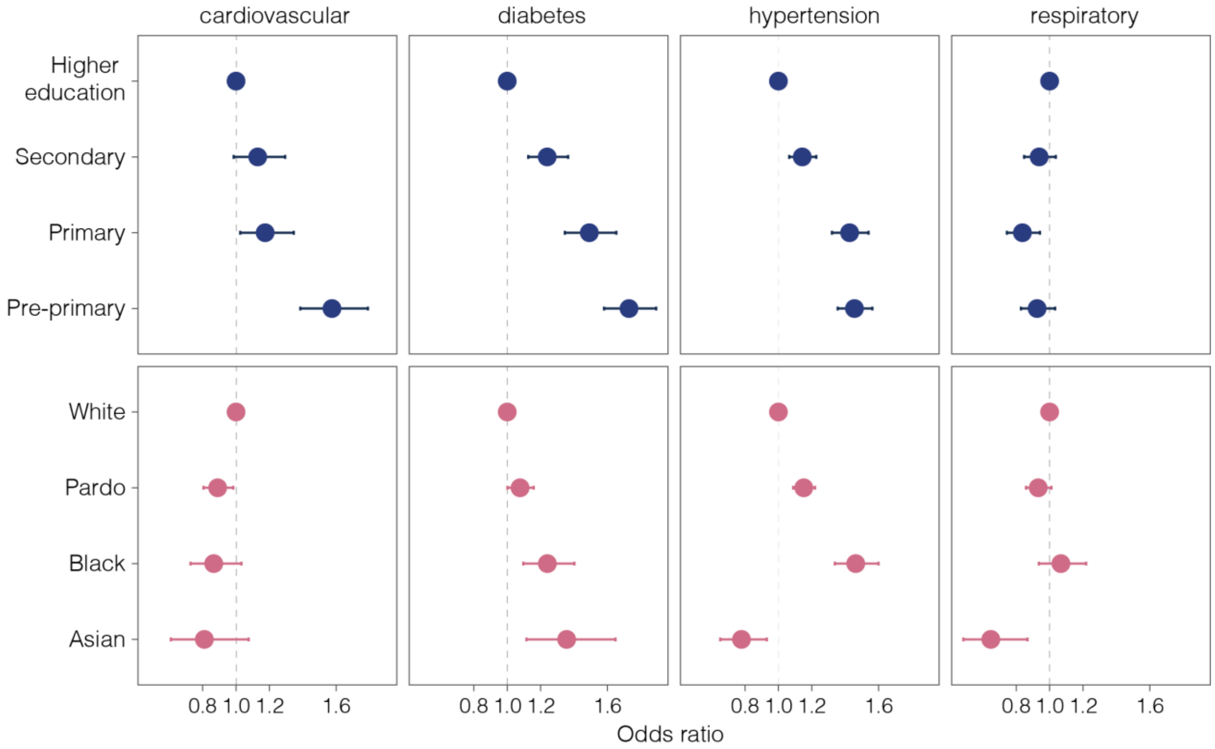

Figure S 4. Odds ratio of having been diagnosed with a comorbidity, by race and education attainment in São Paulo State, 2020. Source: PNAD COVID-19(IBGE).

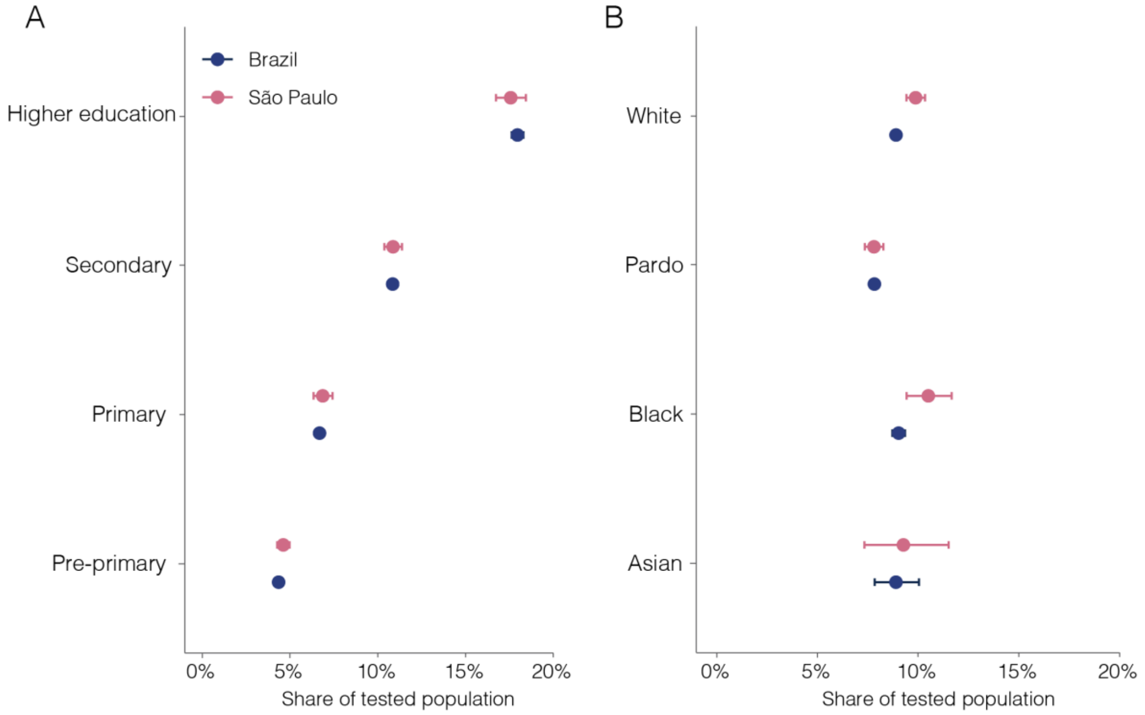

Figure S 5. Proportion of population that reported of being tested for COVID-19 between July and September 2020 by education attainment (A) and race (B) in Brazil and São Paulo state. Source: PNAD COVID-19 (IBGE).

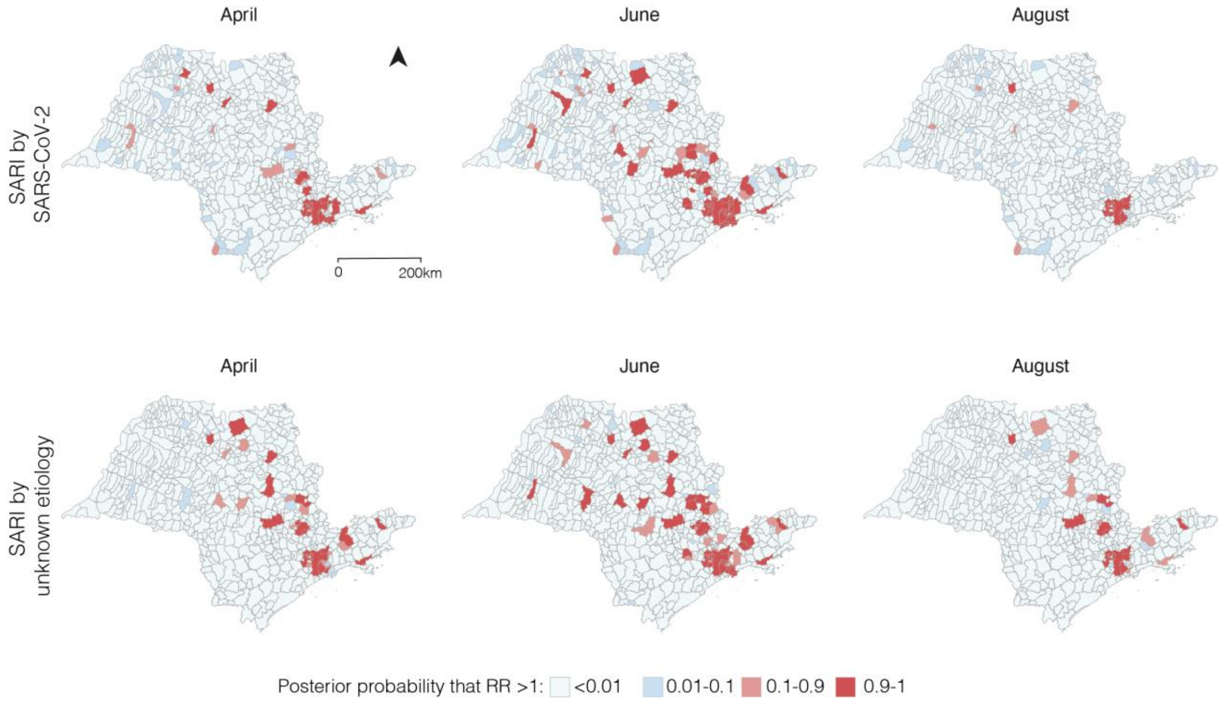

Figure S 6. Posterior probability of elevated relative risk at the municipality level for São Paulo state.
